# Replicated blood-based biomarkers for Myalgic Encephalomyelitis not explicable by inactivity

**DOI:** 10.1101/2024.08.26.24312606

**Authors:** Sjoerd Viktor Beentjes, Julia Kaczmarczyk, Amanda Cassar, Gemma Louise Samms, Nima S. Hejazi, Ava Khamseh, Chris P. Ponting

**Author notes:** Corresponding authors, alphabetical order, equal contribution,.

## Abstract

Myalgic Encephalomyelitis (ME; sometimes referred to as chronic fatigue syndrome [CFS]) is a relatively common and female-biased disease of unknown pathogenesis that profoundly decreases patients’ health-related quality-of-life. ME/CFS diagnosis is hindered by the absence of robustly-defined and specific biomarkers that are easily measured from available sources such as blood, and unaffected by ME/CFS patients’ low level of physical activity. Previous studies of blood biomarkers have not yielded replicated results, perhaps due to low study sample sizes (*n <* 100). Here, we use UK Biobank (UKB) data for up to 1,455 ME/CFS cases and 131,303 population controls to discover hundreds of molecular and cellular blood traits that differ significantly between cases and controls. Importantly, 116 of these traits are replicated, as they are significant for both female and male cohorts. Our analysis used semi-parametric efficient estimators, an initial Super Learner fit followed by a one-step correction, three types of mediators, and natural direct and indirect estimands, to decompose the average effect of ME/CFS status on molecular and cellular traits. Strikingly, these trait differences cannot be explained by ME/CFS cases’ restricted activity. Of 3,237 traits considered, ME/CFS status had a significant effect on only one, via the “Duration of walk” (UKB field 874) mediator. By contrast, ME/CFS status had a significant direct effect on 290 traits (9%). As expected, these effects became more significant with increased stringency of case and control definition. Significant female and male traits were indicative of chronic inflammation, insulin resistance and liver disease. Individually, significant effects on blood traits, however, were not sufficient to cleanly distinguish cases from controls. Nevertheless, their large number, lack of sex-bias, and strong significance, despite the ‘healthy volunteer’ selection bias of UKB participants, keep alive the future ambition of a blood-based biomarker panel for accurate ME/CFS diagnosis.

## 1. Introduction

Myalgic encephalomyelitis (ME; also known as chronic fatigue syndrome, CFS) is an often debilitating disease of unknown pathogenesis defined by post-exertional malaise (PEM), the dramatic worsening of symptoms after even minor mental or physical exertion [1], which usually persists at least 24 hours, in contrast to other fatiguing illnesses [2]. ME/CFS has no cure and no widely effective therapy [3]. It is a female-dominant disease [4, 5]. There are no clinical biomarkers for ME/CFS. A high priority for people with ME/CFS is an accurate and reliable diagnostic test [6]. Findings from dozens of biomarker studies have shown limited reproducibility, perhaps due to their typically low sample sizes, their frequent use of inappropriate statistical tests [7] and the known heterogeneity of ME’s symptoms and potential aetiology [8].

Any clinical biomarker would need to account for individuals’ inactivity relative to the general population. This is because many people with ME/CFS do not exercise and have to restrict their activity [9] to reduce the risk of subsequent PEM. Some have proposed that it is this avoidance of activity that inhibits recovery by perpetuating ME/CFS symptoms following an acute illness [10, 11, 12]. According to this theory, a gradual return to activity reduces fatigue and disability by reversing deconditioning [13] and would reverse any physiological changes, for example in blood traits, caused by inactivity [14]. Counter to this theory, however, is that therapies based on physical activity or exercise are ineffective as a cure [15], implying that ME/CFS is instead an ongoing organic illness [16, 17].

In this study, we analysed data from UK Biobank (UKB), a population cohort of 500,000 individuals aged between 40 and 69 at recruitment linked to diverse phenotypic data [18]. Our 3 groups of analyses were on (i) 31 blood cell and 30 blood biochemistry phenotypes; (ii) 251 NMR-measured metabolites; and, (iii) 2,923 proteins. Specifically, we quantify which blood traits, Nuclear Magnetic Resonance (NMR) metabolomics, and proteomics features are significantly different between ME/CFS cases versus controls controlling for age and sex. The large UK Biobank data sets for ME/CFS cases and controls provided substantial statistical power to evaluate hypotheses, also allowing comparison between male-only and female-only analyses, something that had not been previously achievable. We take advantage of three mediators of sedentary lifestyle to determine whether any molecular or cellular trait associated with ME/CFS cases is explicable by physical inactivity.

## 2. Results

### 2.1. Study population: cases and controls

We first defined 1,455 ME/CFS cases and 131,303 non-ME/CFS population control individuals from the UKB [18]. Cases reported their clinical diagnosis of CFS or ME/CFS at least once and poor or fair overall health; controls had no evidence for a ME/CFS diagnosis and were of good or excellent health (see Materials and Methods). For each group of analyses, cases and controls were restricted to those with measurements of 31 blood count and 30 blood biochemistry markers, or 251 NMR metabolites, or 2,923 protein levels, respectively. Collection of these biological samples was contemporaneous with self-reporting of CFS at the first visit to a UKB Assessment Centre (2006-2010). Numbers of samples in each category are shown in Table 1. ME sample sizes for measured outcome in blood traits, NMR metabolites and proteins are shown in Supplementary Fig. S1.

**Table 1.** Numbers of UKB ME/CFS cases or non-ME/CFS controls per category.

|  | Blood traits |  | NMR metabolites |  | Proteomics |  |
| --- | --- | --- | --- | --- | --- | --- |
|  | Cases | Controls | Cases | Controls | Cases | Controls |
| Female | 1,069 | 75,731 | 615 | 41,436 | 126 | 7,963 |
| Male | 386 | 55,572 | 213 | 30,921 | 45 | 5,920 |
| Combined | 1,455 | 131,303 | 828 | 72,357 | 171 | 13,883 |

### 2.2. Molecular and cellular traits significantly associated with ME/CFS

We simultaneously quantified two effects of ME/CFS case status on molecular and cellular traits, the natural direct effect (NDE) and natural indirect effect (NIE) (Fig. 1A). Each measures the difference between averages for ME/CFS cases or controls, weighted by activity level and also correcting for age and sex because levels of some molecules are age-(*e.g.* HRG protein) and/or sex-dependent (*e.g.* ALT). NDE and NIE account for the activity mediator by decomposing the average effect of ME/CFS case status on molecular or cellular trait into (a) direct paths – those not involving the mediator (Fig. 1A, green) – and (b) indirect paths – those acting through the mediator (blue) – with level of activity as the mediator variable [20, 21] (see Materials and Methods).

**Figure 1.**
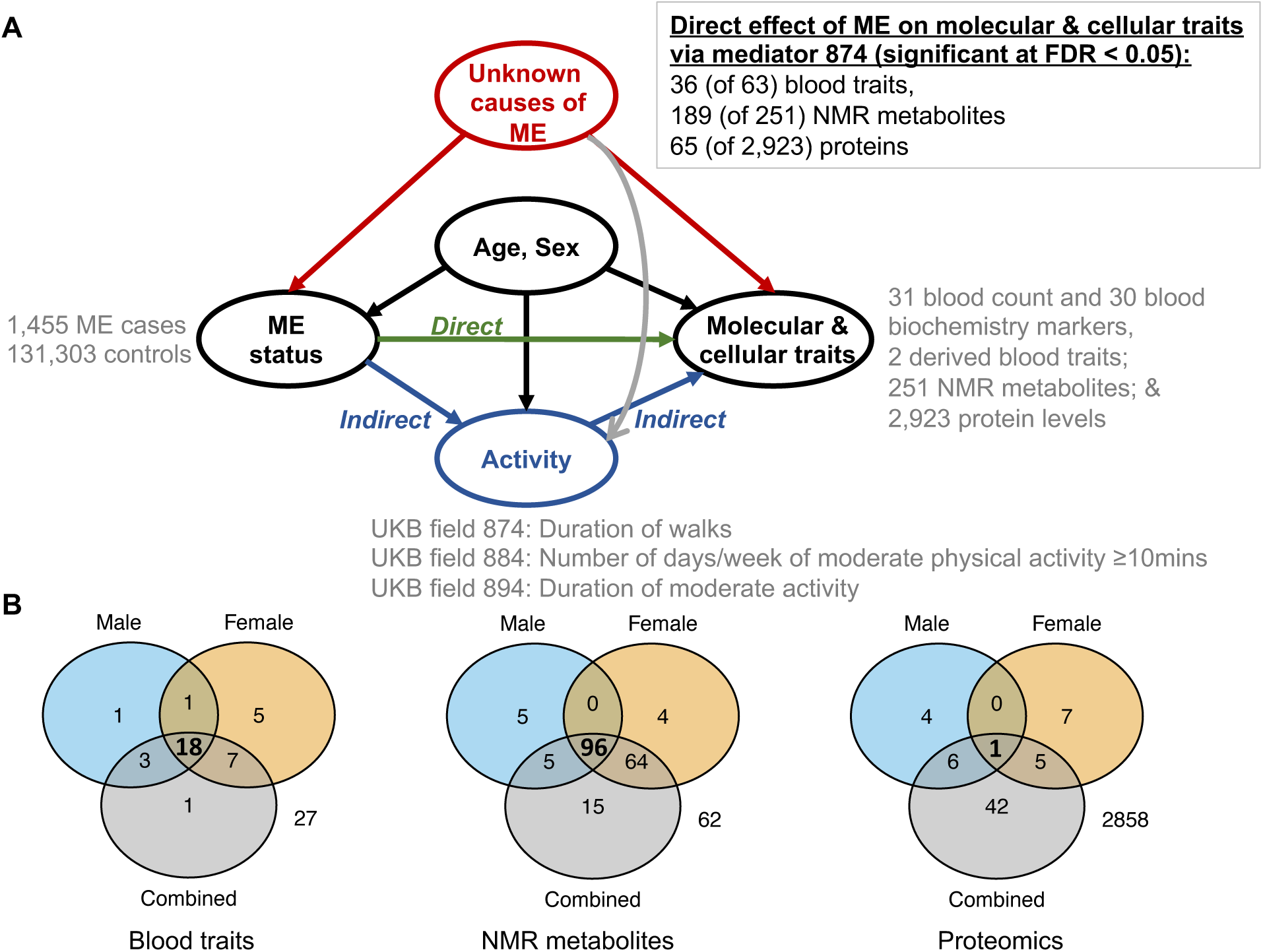
(A) Directed Acyclic Graph for ME/CFS, taking age and sex as confounders and sedentary lifestyle (physical activity) as a mediator for ME/CFS’s effect on molecular and cellular traits. The causes of ME/CFS are an unknown variable (red). Therefore, all effect estimators are quantifying an association between ME/CFS and molecular or cellular traits and no causal statements are made. The “Age” variable (UKB field 21022) represents age at recruitment to UKB, rather than age of onset or diagnosis of ME/CFS. This variable affects the probability of having a ME/CFS diagnosis: recovery is minimal (*≈* 5%, [19]), and as they age people are increasingly likely to be diagnosed with ME/CFS. As it also affects the molecular and cellular traits, age is treated as a confounder. (B) Venn diagrams displaying the number of significant findings in the male, female, and combined cohorts, and their intersection for NDE, mediator 874. Proteomics data have the smallest sample size (see Table 1) and least power, implying fewer significant results in males and females separately as compared to the combined analysis.

As a mediator variable, we first used “Duration of walk” (UKB field 874). As expected, ME/CFS cases reported a lower duration of walk (mean: 44.0 mins/day) than controls (55.3 mins/day). At a false discovery rate [22] (FDR) < 0.05, significant direct effects were found for 36 (of 61 + 2 composite) blood traits, 189 (of 251) NMR metabolites and 65 (of 2,923) proteins (Fig. 1A). All estimates restrict to complete cases, removing individuals with missing trait data in that estimate. For all three analyses, the number of significant NDE results and their intersection in each of the male, female and combined categories, are presented in Fig. 1B.

Significant NDEs on molecular and cellular blood traits for females or males or combined are shown in Fig. 2A. NDEs are strongly correlated between females and males (Fig. 2B). Twenty traits are separately significant in the two sexes (Fig. 2A, B) and thus their associations to ME/CFS status are independently replicated. Additionally, a single trait (erythrocyte_ distribution_width, sometimes a sign of anaemia) was also significant with positive NDE for males and negative NDE for females (Fig. 2A).

**Figure 2.**
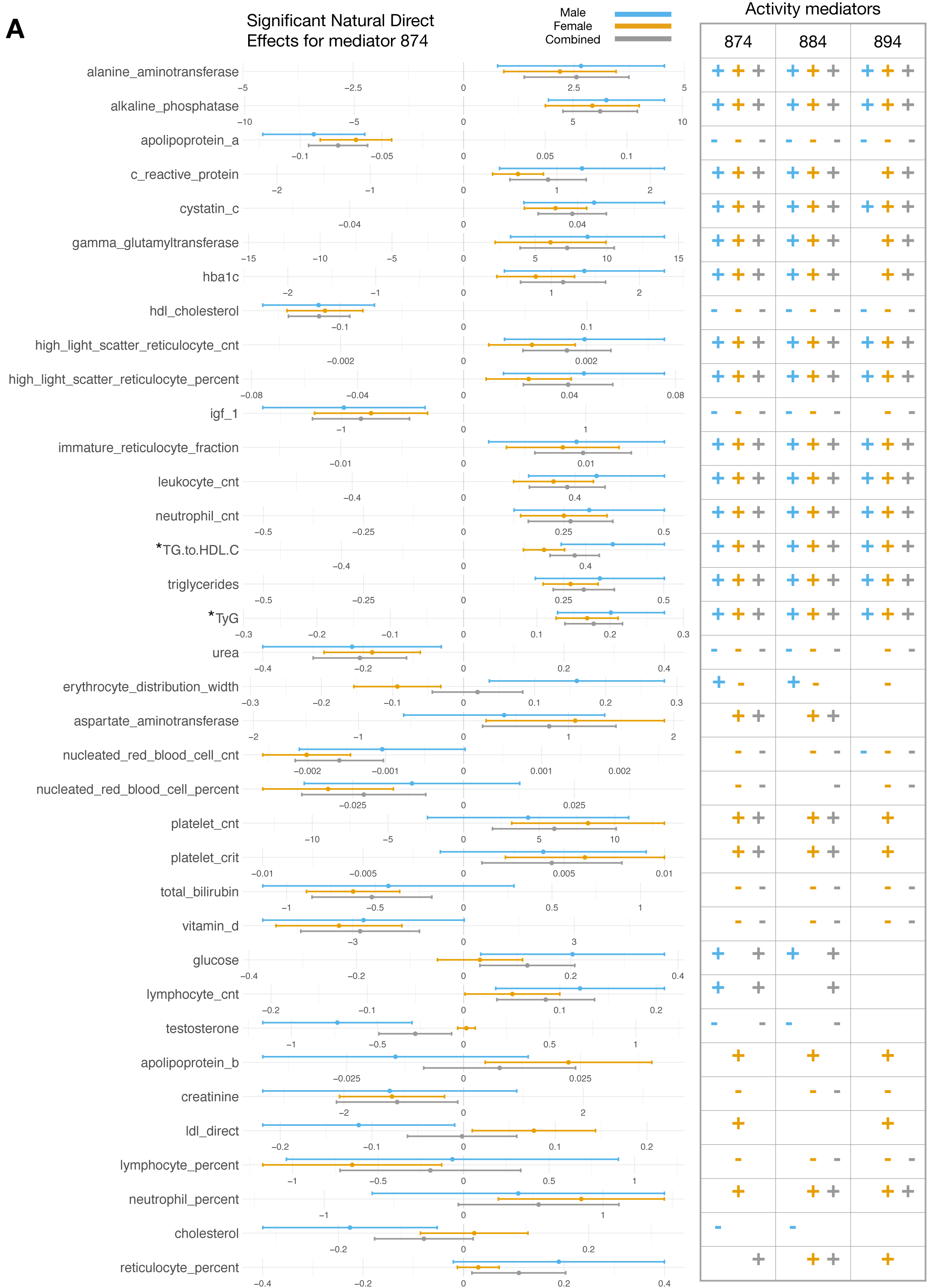
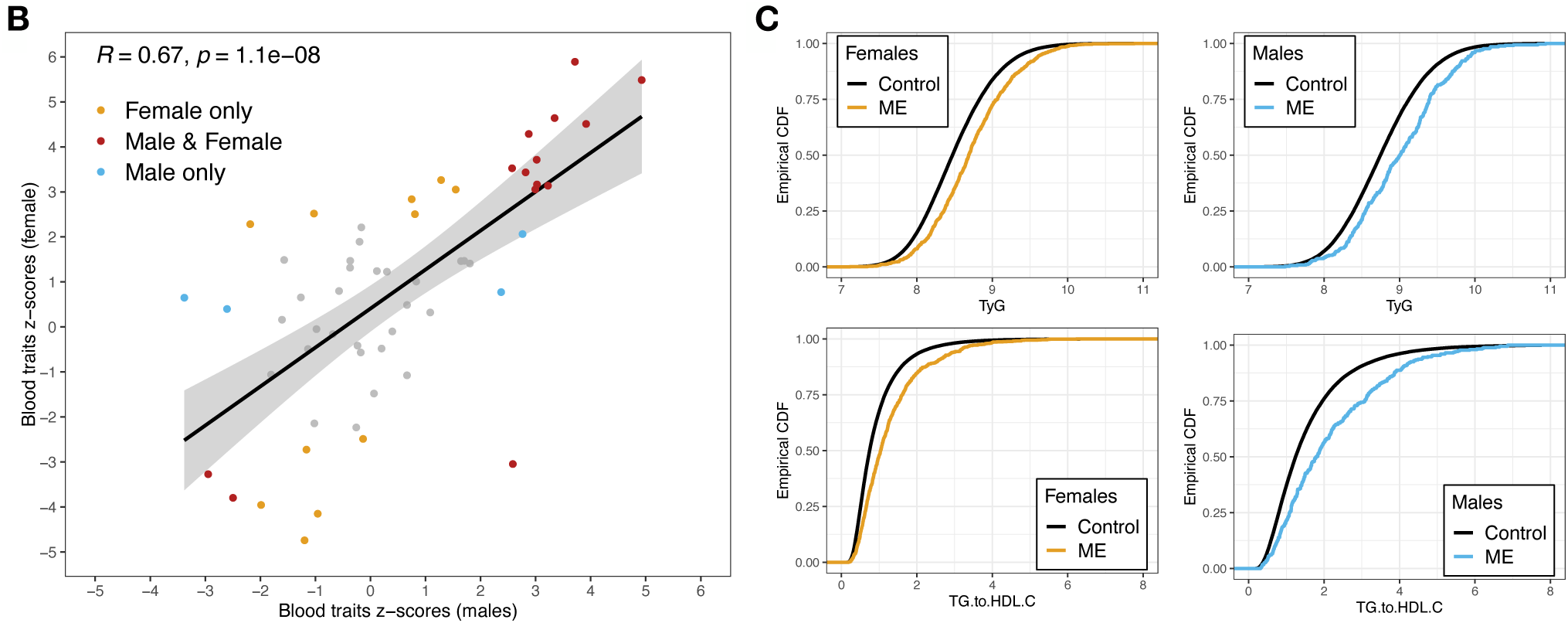
Associational natural direct effects (NDE) of ME/CFS on molecular and cellular blood traits. (A) The sex-stratified analyses are presented in orange (female) and blue (male). For the combined analysis (grey), sex is additionally taken as a confounder. All traits that are significant for the UKB 874 mediator are shown (see Supplementary Table 7 for the UKB 884 and 894 mediators). Natural direct effect sizes (left) are plotted for the UKB 874 mediator (“Duration of walks”), for significant estimates (FDR < 0.05). Error bars indicate 95% confidence intervals. Note that the scale and unit of measurement for each trait (x-axis) are different. For example, the unit of measurement of alanine_aminotransferase (Field 30620) is U/L. The analysis was repeated for the UKB 884 mediator (“Number of days/week of moderate physical activity”) and for the UKB 894 mediator (“Duration of moderate activity”), with the significant results (FDR < 0.05) in each category indicated by ‘+’ symbols for positive effects and ‘*−*’ for negative effects (right). Where there is no symbol, the effect was not significant. Notably, there were no discordant results across the three mediators. All blood trait names are as they appear in the UKB showcase, aside from TyG and TG-to-HDL-C ratio (indicated by *), which are composite measures of other blood traits. (B) Blood trait NDE z-scores, males (x-axis), females (y-axis). Z-scores are the NDE divided by its estimation error. The Pearson correlation is 0.67 and significant. The red dots represent 14 blood traits that are significant in both males and females (FDR < 0.05). The yellow and blue dots represent blood traits that are significant in females only and males only, respectively (FDR < 0.05). The grey dots are significant in neither group while controlling FDR < 0.05. (C) Raw data empirical cumulative distribution functions (ECDFs) for TyG (top) and TG-to-HDL-C ratio (bottom), comparing controls (black) and cases (female on the left, male on the right).

Among the 20 significantly associated traits for females and for males were traits indicative of chronic inflammation (elevated C-reactive protein [CRP] and cystatin C levels, and leukocyte and neutrophil counts), insulin resistance (elevated triglycerides-to-HDL cholesterol [TG-to-HDL-C] ratio, alanine aminotransferase [ALT], alkaline phosphatase [ALP] and gamma glutamyltransferase [GGT]), and liver disease (elevated ALT, ALP and GGT, and low urea levels) (Fig. 2A). Fig. 2C illustrates the shifts in two measures of insulin resistance, the TyG index [23, 24] (top) and TG-to-HDL-C ratio [25] (bottom), between ME/CFS cases and controls. These are the UKB raw data, rather than results from mediation analysis.

### 2.3. Physical activity does not explain ME/CFS-associations to blood traits

Strikingly, for the UKB 874 mediator, significant effects on ME/CFS case status were abundant for direct effects (*i.e.*, NDE; Fig. 2A; Fig. S2), but occurred only once (mean_corpuscular_haemoglobin; adjusted *p* = 0.043) for indirect effects (*i.e.*, NIE; Fig. 3). For every other one of the 61 + 2 composite blood traits, for females or males or both sexes combined, none was significant when controlling the FDR at *≤* 0.05 (Fig. 3). Results from applying two other mediators (Fig. 2A and Fig. 3) are presented later.

**Figure 3.**
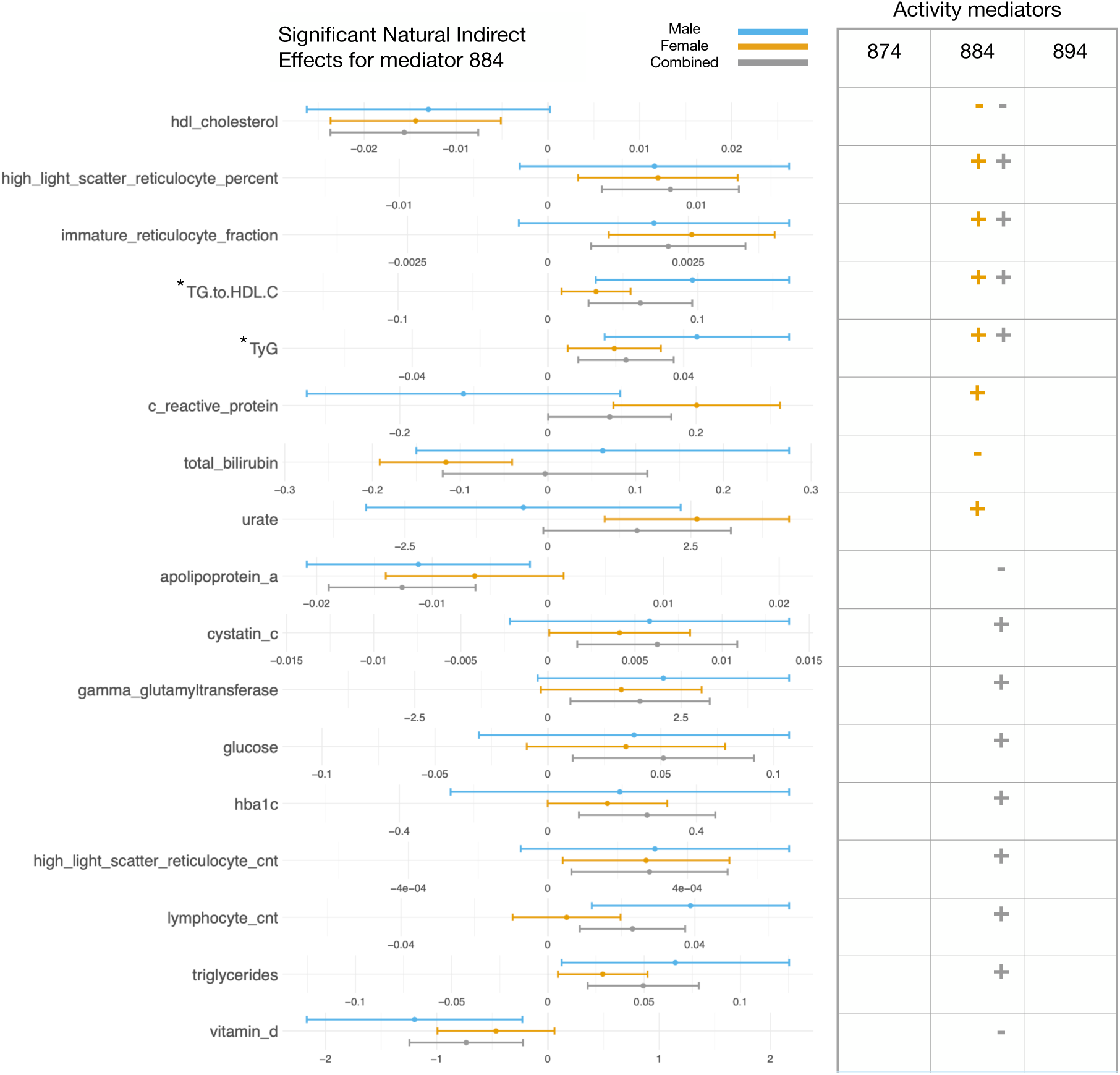
Associational natural indirect effects (NIE) of ME/CFS on molecular and cellular blood traits. The sex-stratified analyses are presented in orange (female) and blue (male). For the combined analysis (grey), sex is additionally taken as a confounder. All traits that are significant for UKB mediator 884 are shown. This is the mediator with the most number of significant indirect effects. UKB mediator 874 “Duration of walks” has a single significant NIE (mean_corpuscular_haemoglobin for females) (FDR < 0.05), whereas UKB mediator 894 “Duration of moderate activity” has no significant NIEs (FDR < 0.05). Effect sizes are plotted for UKB mediator 884 “Number of days/week of moderate physical activity”, for significant estimates (FDR < 0.05). Error bars indicate 95% confidence intervals. Note that the scale and unit of measurement for each trait (x-axis) are different. Significant results (FDR < 0.05) for mediator 884 are indicated by ‘+’ for positive effects and ‘*−*’ for negative effects. Where there is no symbol, the effect was not significant. Blood trait names are as they appear in the UKB showcase, aside from TyG and TG-to-HDL-C ratio (indicated with *) which are composite measures of other blood traits.

### 2.4. Metabolite traits significantly associated with ME/CFS

Of 251 NMR metabolite traits, 189 (75%) were significantly associated with ME/CFS status in an NDE analysis with females only (68 traits) or males only (10 traits) or in both the females only and males only analyses (96 traits) (UKB 874 mediator; Fig. 1B, Supplementary Table 4). Significant traits were mostly lipid levels, involving lipoproteins, cholesterol, and triglycerides. Results were highly concordant between females only and males only analyses (Fig. 4A and B) indicating that ME/CFS-specific blood metabolite differences are, again, generally not sex-biased. Previous ME/CFS metabolomic biomarker studies used one and three orders-of-magnitude fewer cases and controls, respectively [7]. The largest among these identified lowered phosphatidylcholines and cholines in blood from ME/CFS cases ([26], see also [27]), results that we replicated here (Fig. 4A). Higher triglycerides and lower HDL cholesterol in ME/CFS cases, observed using UKB enzymatic assays (Fig. 2A), were also observed as significant in the NMR metabolomics assays (Fig. 4A). Of 9 amino acids measured, only alanine was significantly elevated, and then only in female ME/CFS cases. Blood pyruvate and lactate, previously predicted to be ME/CFS biomarkers [28, 29], were also not significantly different between cases and controls.

**Figure 4.**
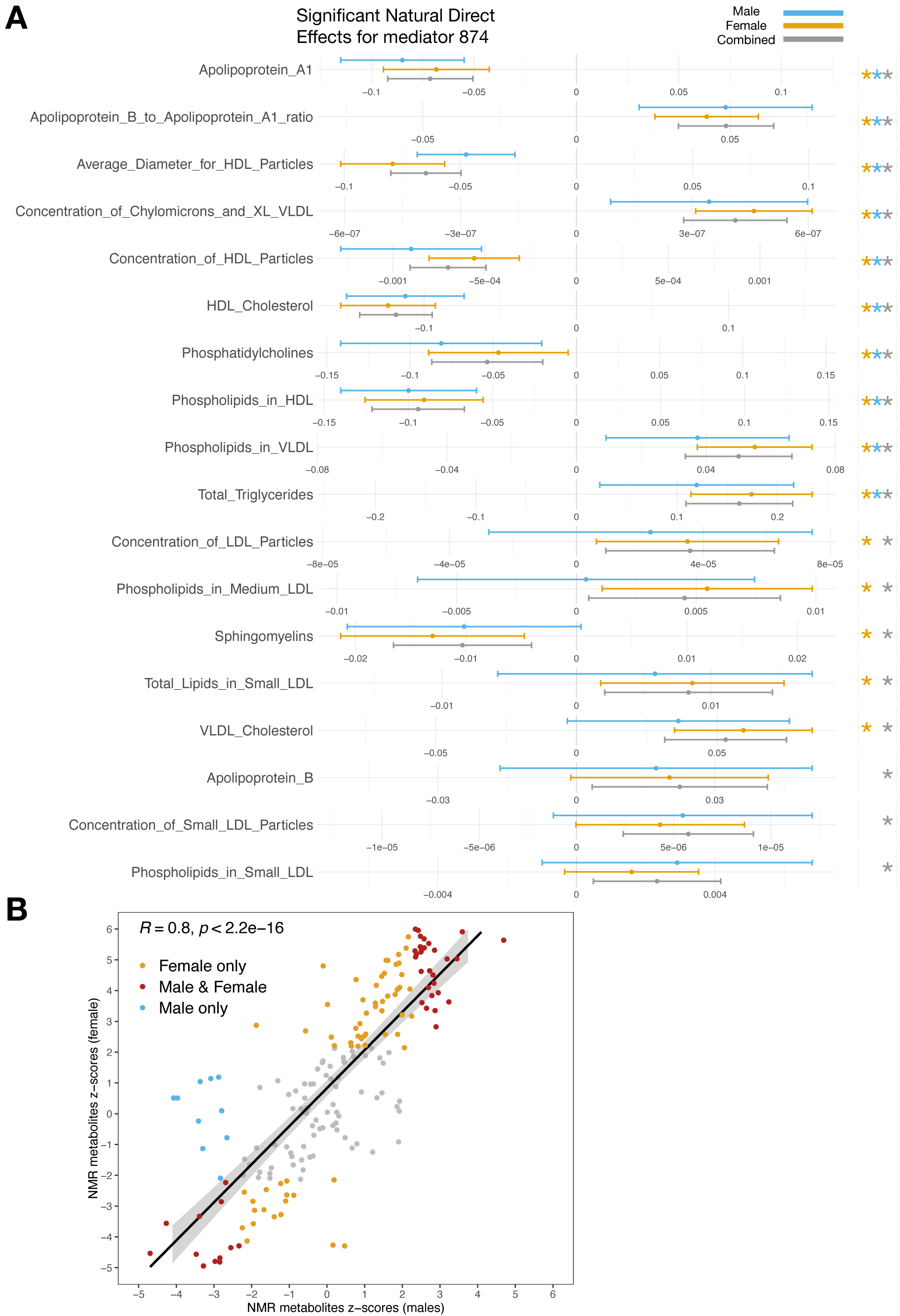
Associational natural direct effects (NDE) of ME/CFS on NMR metabolites. (A) The sex-stratified analyses are presented in orange (female) and blue (male). For the combined analysis (grey), sex is additionally taken as a confounder. Eighteen of 184 traits are shown; results for all traits are provided in Supplementary Table 4. Effect sizes are plotted for mediator 874 “Duration of walks” for significant estimates (FDR < 0.05). Error bars indicate 95% confidence intervals. Note that the scale and unit of measurement (X-axis) are different for each metabolite. Asterisks (right) indicate effects that are significant (FDR < 0.05). Where there is no asterisk, the effect was not significant. There were no discordant results across the three analyses. All NMR metabolite names are as they appear in the UKB showcase. (B) NMR NDE values are strongly concordant between the two sexes. Shown are per-metabolite z-scores for males (x-axis) and females (y-axis). Z-scores are the NDE divided by its estimation error. The Pearson correlation is 0.8 and significant. Red dots indicate metabolites that are significant in both males and females (FDR < 0.05). Yellow and blue dots represent metabolites that are significant in females only and males only, respectively (FDR < 0.05). Grey dots are significant in neither.

None of the 251 metabolite traits was significant when controlling the FDR at *≤* 0.05 for indirect effects using the “Duration of walk” (UKB 874) mediator, for females or for males or for both combined (Fig. S2B).

### 2.5. Proteomic traits significantly associated with ME/CFS

Repeating this NDE analysis using the UKB 874 mediator on levels of 2,923 proteins, measured using antibody-based assays, yielded only a single protein, extracellular superoxide dismutase or SOD3, whose abundance was significantly altered (FDR < 0.05) between cases and controls in both females and males. Relative to preceding analyses, this proteomic analysis is under-powered owing to there being fewer cases for whom data was available (Table 1) and its larger multiple testing burden. Implications of this association to SOD3 are unclear, although superoxide, SOD3’s substrate, is known to modulate the hyperalgesic response [30].

Male- or female-specific effects for the same protein are again correlated (Fig. 5; Supplementary Table 5). Considering all cases combined, 54 proteins are significant (FDR < 0.05; Figure 1B). Among these are 7 complement proteins (C1RL, C2, CFB, CFH, CFI, CFP and CR2) of the innate immune system, whose levels are all elevated in ME/CFS cases, including CR2 (complement C3d receptor 2), the receptor for Epstein-Barr virus (EBV) binding on B and T lymphocytes. Two of the up-regulated proteins (CDHR2 and CDHR5) together form the extra-cellular portion of the intermicrovillar adhesion complex, whose disruption leads to intestinal dysfunction and inflammatory bowel disease [31, 32]. ME/CFS cases also show increase in levels of leptin (LEP), which has a role in energy homeostasis [33]. Again, not a single protein among the 2, 923 yielded a significant NIE estimate for this mediator (Fig. S2B).

**Figure 5.**
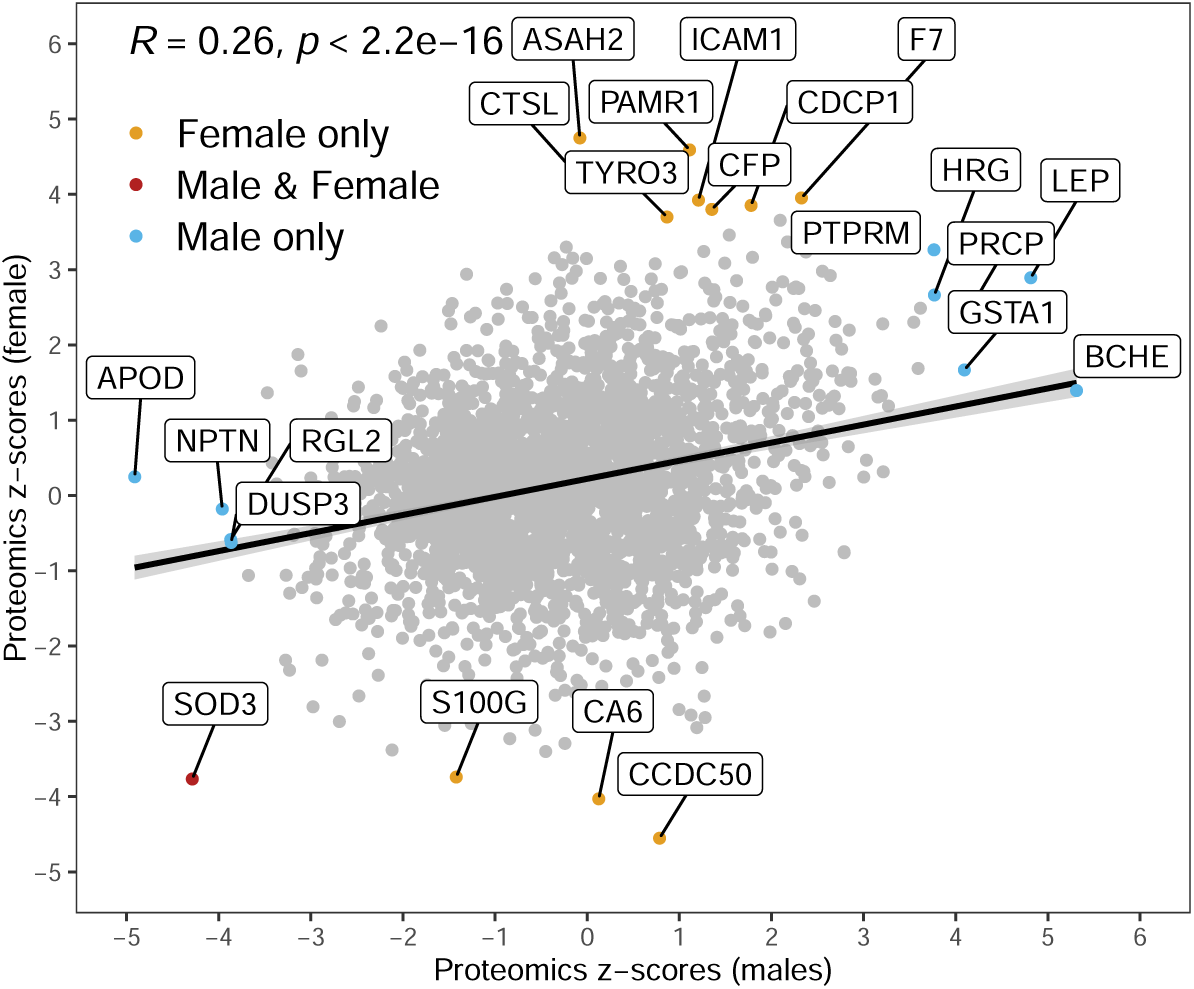
Protein NDE z-scores, males (x-axis), females (y-axis). Z-scores are the NDE divided by its estimation error. The Pearson correlation is 0.26 and significant. The red dot represents the single protein (SOD3) that is significant in both males and females (FDR < 0.05). Yellow and blue dots indicate proteins that are significant in females only and in males only, respectively (FDR < 0.05). Grey dots show proteins that are significant in neither (*i.e.*, FDR *≥* 0.05).

**Figure 6.**
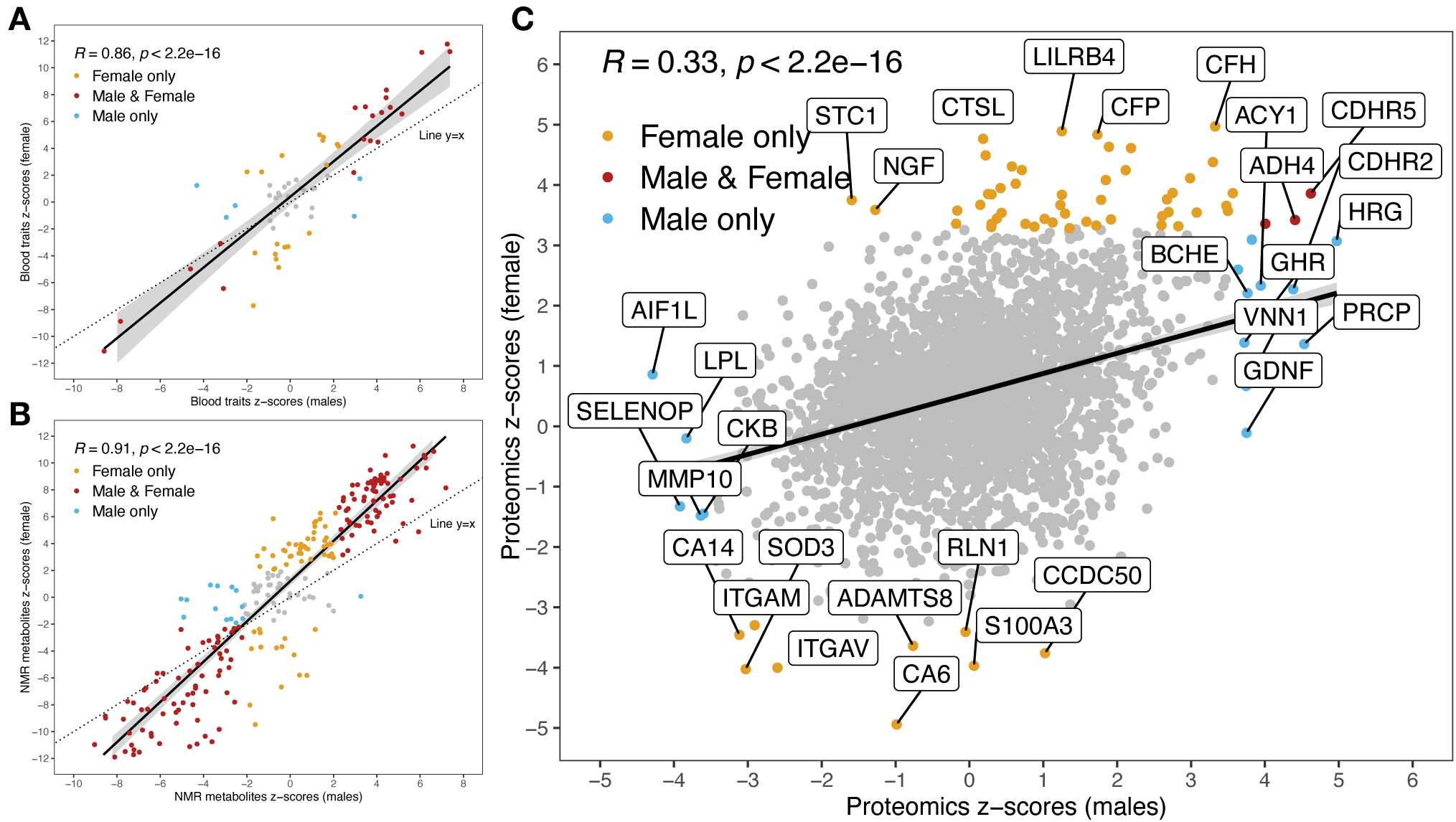
(A) Blood trait total effect z-scores, males (x-axis), females (y-axis). Z-scores are the TE divided by its estimation error. The Pearson correlation is 0.86 and significant. The red dots represent 20 blood traits that are significant in both males and females (FDR < 0.05). The yellow and blue dots represent blood traits that are significant in females only and males only respectively (FDR < 0.05). The grey dots are significant in neither for FDR < 0.05. The *x* = *y* line indicates the line of equal z-scores for males and females. In general, in absolute value, the z-scores are higher for females than males. This is to be expected as the sample size is larger for females. (B) Metabolite total effect values are strongly concordant between the two sexes. Shown are per-metabolite z-scores for males (x-axis) and females (y-axis). Z-scores are the TE divided by its estimation error. The Pearson correlation is 0.91 and significant. Red dots indicate metabolites that are significant in both males and females (FDR < 0.05). Yellow and blue dots represent metabolites that are significant in females only and males only, respectively (FDR < 0.05). Grey dots are significant in neither. (C) Proteins total effect z-scores, males (x-axis), females (y-axis). Z-scores are the TE divided by its estimation error. The Pearson correlation is 0.33 and significant. Red dot represents the proteins (LEP, CDHR5, ADH4, RTN4R) that are significant in both males and females (FDR < 0.05). Yellow and blue dots represent proteins that are significant in females only and males only respectively (FDR < 0.05). The grey dots are significant in neither for FDR < 0.05.

### 2.6. Total effects

We have shown above that direct effects dominate, so that indirect effects contribute little-or-nothing to molecular and cellular effects. In real-world settings, the quantity of most interest to clinicians will be the total effect (TE), accounting for age and sex, rather than the direct effect. Estimating the total effect for 63 blood traits finds 39 to be significant (FDR < 0.05) predictors of ME/CFS case status for females and males combined (Supplementary Fig. S2A). The traits that are robustly predictive of ME/CFS are those shown in Fig. 2A (with 4 exceptions: erythrocyte_distribution_width, apolipoprotein_b, creatinine and ldl_direct). For one or more of female- or male-specific or combined TE analyses, a total of 251 proteins and 216 metabolites were additionally significant (FDR < 0.05; Supplementary Fig. S2A).

Significantly enriched Gene Ontology (GO) terms for TE-significant proteins highlighted tumour necrosis factor (TNF) and interleukin-4 (IL4) production, and natural killer (NK) cell mediated cytotoxicity (Fig. S3). Nevertheless, TNF and IL4 proteins themselves were not significantly altered in abundance. Impaired NK cell cytotoxicity in ME/CFS, however, is one of the few cellular or molecular biomarkers that has often been replicated [34].

### 2.7. Sensitivity analyses for blood traits

Next, we investigated whether blood trait results replicate for 2 further mediators: “Number of days/week of moderate physical activity 10+ mins” (UKB field 884) and “Duration of moderate activity” (UKB field 894) questionnaire responses. As expected, ME/CFS cases reported less activity than controls: mean 2.77 vs 3.51 days/week, and 53.9 vs 60.0 mins/day for mediators 884 and 894, respectively. As before, significant effects on ME/CFS status were observed for direct effects, never indirect effects for the “Duration of moderate activity” mediator (UKB field 894) (Fig. 2A, Fig. 4). By contrast, for the “Number of days/week of moderate physical activity 10+ mins” (UKB field 884) mediator, 22 significant NIEs were identified: 14 (0.4%) and 8 (0.2%) traits at FDR *≤* 0.05 for combined female and male, and female-only data, respectively. Importantly, even when significant NIEs are found, they almost always contribute less to the total effect than NDEs (Fig. 7).

**Figure 7.**
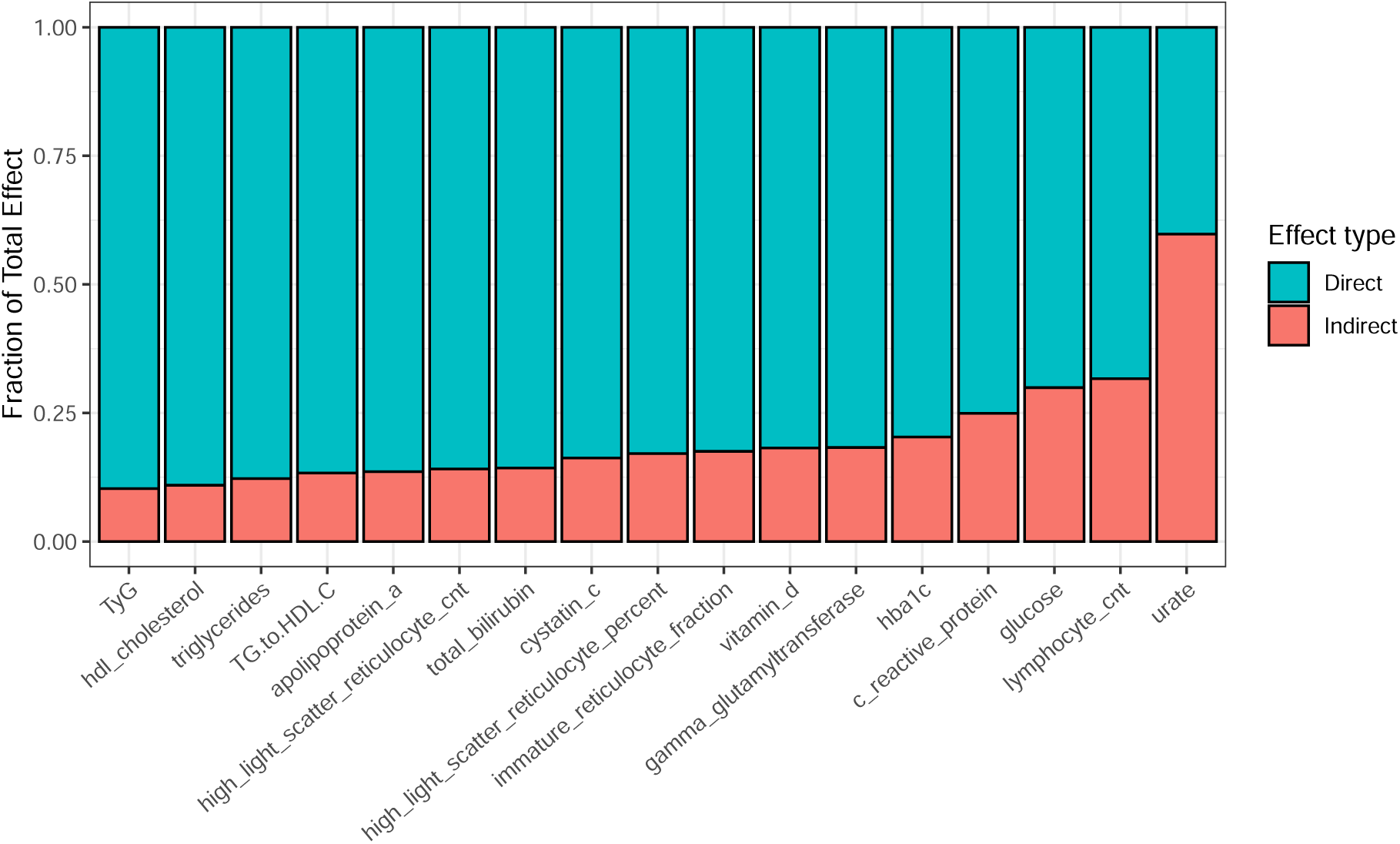
Associational NDE (blue) and NIE (red) as a fraction of the total effect for the effect of ME/CFS on molecular and cellular blood traits. The results are presented for male and female combined, for mediator 884 “Number of days/week of moderate physical activity”, the only mediator that exhibit indirect effects. Across all 61 blood traits, and the two composite metrics TyG and TG-to-HLD-C ratio, only 1 feature, Urate, has a larger NIE than NDE, for this mediator only.

We additionally investigated the dependence of results on the choice of fitting algorithm(s) for blood traits. Specifically, the result in Fig. 2A are obtained using a cross-validating library of algorithms (Super Learner (SL), see Material and Methods). Results obtained with no SL – reducing the library to the baseline GLMnet – with mediator 874, for TE, NDE and NIE are provided in Supplementary Table 9. Although we recommend its use, leaving out the SL has only minor effect: 36 of 39 significant TE blood traits using UKB field 874 as mediator with the SL were also significant without its use, Supplementary Table 9. Full NDE and NIE values for all mediators with SL are provided in Supplementary Table 7.

For TEs, 41 blood traits (as well as TyG and TG-to-HDL-C ratio) differ significantly between female or male ME/CFS cases and controls (Supplementary Table 3). To test whether extreme values affect these results, we winsorized the blood trait data at 0.5% and 1%. The results on the combined dataset are presented in Supplementary Fig. S4 and Supplementary Table 8, and remain robust. To obtain a high confidence set, we further restricted these traits to those significant for NDE for females and for males (mediators 874 and 884) and for females (mediator 894), resulting in 18 traits listed in Supplementary Table 2.

Lastly, we found that TEs and NDEs increase as the stringency of case and control definitions increases (Fig. 8; see Supplementary Table 10 for full results). Specifically, we compared NDEs for molecular and cellular blood traits calculated from cases and controls as defined in Materials and Methods, but with or without overall health rating (UKB field 2178) of ‘Poor’ or ‘Fair’ at baseline for cases, and/or ‘Good’ or ‘Excellent’ for controls. Removing general health criteria when defining cases and controls did not substantially affect our statistically-significant discoveries (Fig. 8).

**Figure 8.**
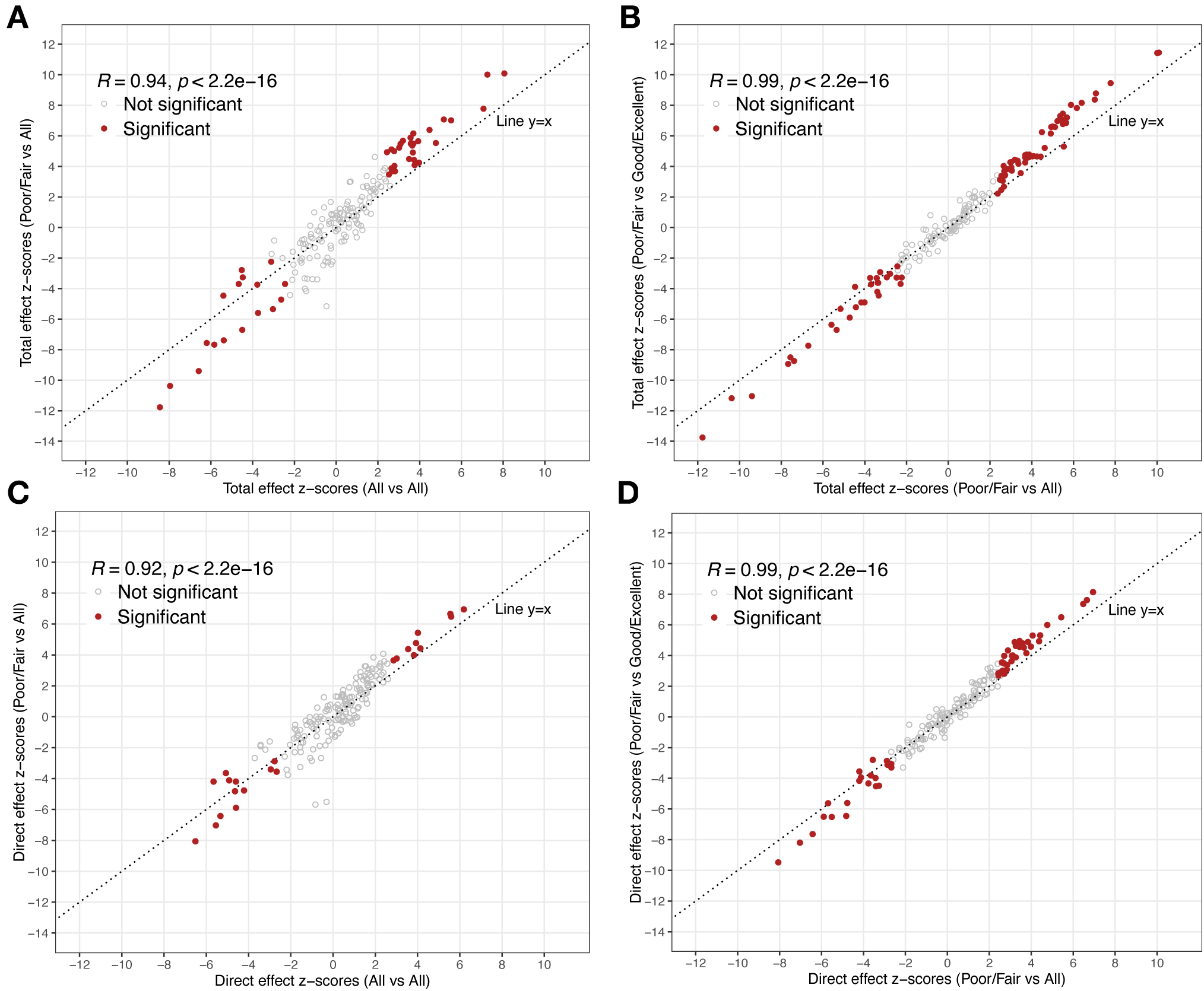
Total effects and NDEs for blood traits become more significant as the stringency of case and control definitions increases. (A) Total effect z-scores for ‘Poor/Fair’ for cases and ‘All’ (without restricting by health rating (UKB field 2178)) for controls versus z-scores for ‘All’ for cases and ‘All’ for controls (without restricting by health rating for cases or controls). The null hypothesis – that significance does not change for increasing stringency of case or control definition – is represented by the diagonal line. (B) Total effect z-scores for ‘Poor/Fair’ for cases and ‘Good/Excellent’ for controls, versus ‘Poor/Fair’ for cases vs ‘All’ for controls. (C) and (D) As in (A) or (B) but for NDE.

## 3. Discussion

Our results reveal 511 blood-based biomarkers whose levels differ significantly between people with ME/CFS and those without ME/CFS (Fig. S2A). Our approach decomposed the total effect of ME/CFS on blood traits into two components: (1) the indirect effect of ME/CFS on these traits via activity, and (2) the direct effect through all other paths, not mediated via activity. We do not claim causality for our estimates, because the assumptions of no unmeasured confounding may be violated. Nevertheless, any “causal gap”, the difference between our estimates and any underlying causal estimand, cannot be due to age and sex, as we account for these factors. Our findings constitute differences of population estimates of blood biomarkers between case and control populations and do not provide individual-level predictions of caseness based on biomarker values. However, our results can be used for variable selection in training a prediction model, as long as an independent data set is used. If the same data is used twice, *i.e.*, both for variable selection and for training a prediction model, the resulting predictions will suffer from selective inference [35], with overly optimistic (invalid) prediction scores, and thus will not generalise to new cases.

The large number of discoveries relative to previous studies likely reflects our study’s substantially higher numbers of cases and controls (Table 1). These large numbers allow many small average effects of ME/CFS status on molecular and cellular traits to be detected. Importantly, and unlike most previous studies, we independently replicated 166 biomarkers in both females and males (TEs; Fig. S2A). This indicates that our discoveries are both robust and not sex-biased. It thus provides strong evidence for ME/CFS disease pathophysiology being equivalent in both sexes. This is despite sex-bias of ME/CFS with respect to prevalence and onset, comorbidities, symptoms and other features [4, 36].

Importantly, these biomarker differences are not explicable by dissimilarities in physical activity: among 3,237 NIE estimates we obtained, ME/CFS status was significantly associated with only one trait (Fig. S2B). Blood traits thus distinguish ME/CFS cases from population controls, but not because of ME/CFS cases’ reduced physical activity levels.

What then cause these molecular and cellular changes in blood if not physical activity? Our findings provide strong and replicated evidence for chronic low-level inflammation (elevated CRP and cystatin-C levels, and platelet, leukocyte and neutrophil counts), insulin resistance (elevated triglycerides-to-HDL-C ratio, ALT, ALP, GGT and HbA1c) and/or liver disease (elevated ALT, ALP, and GGT, and low urea levels) in ME/CFS (Fig. 2A). ME/CFS is thus portrayed by insulin resistance and systemic inflammation, with liver inflammation and dysfunction likely affecting lipid metabolism and the balance between HDL and LDL cholesterol. To our knowledge, the overall combination of blood marker changes we observed does not present in any other disease. For example, although primary biliary cholangitis is accompanied by elevated ALP and GGT levels (and post-exertional malaise-like symptoms [37]) it is also marked by high circulating levels of bilirubin rather than the lower levels we observe for ME/CFS (Fig. 2A). Nevertheless, because ME/CFS likely arises from multiple pathomechanisms and we did not further stratify cases, we cannot conclude that our results exclude other diseases from sharing a common aetiology with some ME/CFS cases.

In general, shifts in trait values were modest. Among all 116 significant female- and male-replicated traits (NDEs), 91% had small-to-medium shifts (Cohen’s *d* between 0.2 and 0.5 [38]; Supplementary Table 11). No trait yielded clear separation in estimated effects between ME/CFS cases and controls, rather trait values overlapped extensively. For example, despite CRP level being significantly elevated in ME/CFS cases (TE analysis: adjusted *p* = 2.8 *×* 10*^−^*^9^; both sexes), only 4.8% of female and 2.5% of male ME/CFS cases (versus 2.2% and 1.8% controls, respectively) had CRP levels over 10mg/L, a moderate elevation that can indicate systemic inflammation in autoimmune disease. Consequently, no single blood trait we analysed will be an effective biomarker for ME.

The major strength of the study is its large and deeply phenotyped cohort who were recruited, and their blood traits measured, using a single protocol. The study also controlled for potential confounders such as age, sex and physical inactivity. Additional mediators beyond physical activity were not considered as they were not directly relevant to this study’s principal hypothesis. The study was limited by the UK Biobank’s known healthy volunteer bias [39], possibly resulting in few, if any, people with severe ME/CFS symptoms at baseline participating. Participants’ diet, medication, smoking, alcohol use and socioeconomic status could be confounders but only if they causally influence ME/CFS status. Future studies could test for the effect of symptom severity on the levels of biomarkers found to be significant in this study. UK Biobank recruited 40-69 year old participants [39], an age range when individuals are less likely to have a clinical diagnosis of ME/CFS [40]. Our analysis relies also on correct clinical categorisation of ME/CFS disease and participants’ recall of it. We note that the list of cellular and molecular measurements in the UK Biobank is not exhaustive. For example, others have investigated potential biomarkers for oxidative stress [41] as well as gut metagenomics, immune-profiling and cytokines [42] which are absent from UKB.

Evidence that there is a large number of replicated and diverse blood biomarkers that differentiate between ME/CFS cases and controls should now dispel any lingering perception that ME/CFS is caused by deconditioning and exercise intolerance [10, 11, 12, 13] These findings should also accelerate research into the minimum panel of blood traits required to accurately diagnose ME/CFS in real-world populations. Such a panel would be invaluable for diagnosis, for measuring response to future treatment or drug trials, and potentially for determining the worsening or progression of ME/CFS. Such a panel might also help to determine the distinctions or overlap between ME/CFS and symptomalogically similar diseases such as Long Covid and fibromyalgia.

To assist the search for an effective biomarker panel for ME/CFS we provide the full results of this study in Supplementary Tables 3-5.

## 4. Materials and Methods

### 4.1. UK Biobank ME/CFS data processing

We defined 1,455 ME/CFS cases and 131,303 non-ME/CFS control individuals from UKB [18] as follows. Cases self-reported a diagnosis of ‘Chronic Fatigue Syndrome’ (CFS) in verbal interview at their first visit to a UKB Assessment Centre (UKB field 20002); also, either they answered “Yes” to the question “Have you ever been told by a doctor that you have Myalgic Encephalomyelitis/Chronic Fatigue Syndrome?” in the ‘Experience of Pain Questionnaire’ (PQ) (2019-2020) (UKB field 120010), or they did not complete the PQ. They further reported an overall health rating (UKB field 2178) of ‘Poor’ or ‘Fair’ at baseline, and were of known genetic sex. Population controls did not self-report a CFS diagnosis in any of the 4 visits, answered “No” to the PQ question about a ME/CFS diagnosis, and were not linked to a Primary Care record (CTV3 or Read v2 code, Supplementary Table 1) of ME/CFS or to the ICD10:G93.3 code (‘Postviral fatigue syndrome’) in Hospital Inpatient Data. They further reported an overall health rating (UKB field 2178) of ‘Good’ or ‘Excellent’ at baseline. UKB participants are older and report healthier lifestyles, higher levels of education and better health relative to the general UK population [43, 44]. UKB assessment at baseline was demanding in time (2-3h) and energy, including travel to the nearest of 22 centres. These requirements will have diminished the recruitment of people with severe or moderate, or even mild, ME/CFS symptoms. UKB blood samples were acquired and analysed as described previously [45, 46, 47]. On average, the body mass index (BMI; UKB field 21001) of cases is significantly, but only slightly, higher than the BMI of controls (27.96 *±* 4.62 for 386 male cases vs 26.80 *±* 3.58 for 55, 572 male controls, with *t* = 4.92 and *p* = 10*^−^*^5^, Welch’s t-test; 27.70 *±* 5.83 for 1069 females cases vs 25.82 *±* 4.39 for 75, 731 female controls with *t* = 10.68 and *p <* 10*^−^*^12^, Welch’s t-test).

For blood traits, we included two composite markers of insulin resistance: the triglyceride glucose (TyG) index [48, 49], and TG-to-HDL-C ratio [50]. Note that TyG is normally calculated using fasting levels of triglycerides and plasma glucose [51], but these are not available from the UK Biobank. The ratio of triglycerides to HDL-cholesterol correlates inversely with the plasma level of small, dense LDL particles.

For NMR metabolomics, we removed individuals whose NMR metabolite measurement has a QC flag indicating irregularities in the measurement, as per UKB category 221.

For each estimator of type TE, NDE and NIE (below), we only considered individuals with the relevant variables measured. Specifically, for TE, we restricted to individuals with measured age, sex and outcome variable. For NDE and NIE, we additionally restricted to individuals with measured mediators of activity. Furthermore, for NDE and NIE, we removed individuals who answered ‘do not know’ or ‘prefer not to answer’ to the activity question (UKB datafield 874, 884, or 894).

### 4.2. Mediation estimators

Causal mediation analysis, concerned with the quantification of the portion of a causal effect of an exposure on an outcome through a particular pathway, has been extensively discussed in the literature [52, 53]. The methodologies utilised in this work build upon natural (or pure) mediation estimands [20, 21]. Strategies for the construction of efficient estimators of non-parametrically defined causal mediation estimands, capable of incorporating machine learning, have been used in a variety of applications. Recent examples include understanding the biological mechanisms by which vaccines causally alter infection risk [54, 55, 56, 57], quantifying the effect of novel pharmacological therapies on substance abuse disorder relapse [58, 59] and the effects of housing vouchers on adolescent development [60], and modeling the effects of health disparities on quality of life [61]. Here we use state-of-the-art semi-parametric estimation techniques for non-parametric causal mediation analysis [62], implemented in the R package medoutcon [63, 64].

The NDE and NIE are mediational estimands that decompose the average effect (or average treatment effect, ATE) of ME/CFS status on molecular and cellular traits, Eq. 1.

NDEs involve a comparison of two counterfactual trait outcomes, specifically:

(I) the level of the trait in a hypothetical scenario where every individual has ME, but rather than allowing ME/CFS to determine the level of activity, we fix their level of activity to the values they would naturally assume if they were not to have ME; and,
(II) the level of the trait in a hypothetical scenario where every individual is in the control group and their levels of activity are allowed to naturally respond to being in the control group.

Comparison of these two trait levels yields a “direct” causal effect that quantifies the effect of ME/CFS on the trait through all paths other than the one mediated by activity.

NIEs involve a comparison of two counterfactual trait outcomes, specifically:

(i) (III) the level of the trait when every individual has ME/CFS and their levels of activity are allowed to naturally respond to ME; and,
(ii) (IV) the level of the trait in a hypothetical scenario where every individual has ME, but rather than allowing ME/CFS to determine the level of activity, we fix their activity level to the value they would naturally assume if they were not to have ME.

Comparison of these two trait levels yields a causal “indirect” effect that quantifies the impact of ME/CFS on trait through activity (NIE). Crucially, the counterfactual trait outcomes (I) and

(i) (IV) are exactly the same quantity, and this insight gives rise to the “mediation formula” as follows:

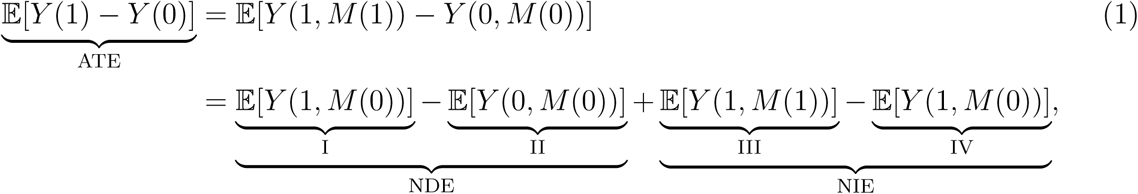

where *Y* (1) and *Y* (0) are potential outcomes in which an individual does or does not have ME, respectively. Similarly, *Y* (1, *M* (0)) is the potential outcome of an individual who has ME/CFS and whose mediator takes on the value it would have had if the individual did not have ME/CFS (given in words as (I) and (IV) above). Note also that *Y* (1) = *Y* (1, *M* (1)) and *Y* (0) = *Y* (0, *M* (0)). The left hand side of Eq. 1 defines the average treatment effect (ATE) of ME/CFS on blood trait *Y*, which we refer to as the total effect (TE). The right hand side of this equation is the sum of the NDE and NIE.

Causal identification is the process of turning a causal quantity we wish to estimate (causal estimand – a functional of unobservable counterfactual data) into a statistical quantity we can estimate from observed data (statistical estimand – a functional of observed data). Causal identification does not require access to any data and is entirely distinct from statistical inference. There are 5 assumptions required for causal identifiability of Eq. 1:

(i) the Stable Unit Treatment Values Assumption (SUTVA) which includes consistency and no interference between units [65, 66];
(ii) exchangeability (unconfoundedness), which is analogous to the randomization assumption applied to a joint intervention on both the treatment variable (here ME/CFS) and the mediator (here activity);
(iii) treatment positivity, which states that it must be possible to observe any given treatment value (here ME/CFS) across all strata of baseline covariates (age and sex);
(iv) mediator positivity, which states that it must be possible to observe any given mediator value across all strata defined by both treatment (ME/CFS) and baseline covariates (age and sex); and,
(v) Cross-world counterfactual independence *Y* (*T* = *t, M* = *m*) *⊥⊥ M* (*T* = *t^′^*) conditional on covariates, which is not empirically verifiable [67].

In our case, we do not claim causal identifiability because the assumptions of unconfoundedness (ii) may be violated, as made explicit in Fig. 1A (in red). Nevertheless, we can estimate the NDE and NIE as statistical quantities knowing that any causal gap will not be due to age or sex, as both of these variables have been taken into account as confounders.

### 4.3. Super Learner and one-step estimation

We have used semi-parametric efficient estimators to estimate the TE, as well as the mediation effects NDE and NIE [64], on multiomic measurements. This estimation procedure consists of an initial Super Learner (SL) [68] fit to estimate relevant nuisance functions in as flexible a manner as allowed by the available data. This ensures that any model mis-specification bias is minimised. We then construct estimates of the NDE and NIE using a one-step bias-correction procedure, which appropriately handles the use of SL for nuisance parameter estimation while also allowing for uncertainty quantification, facilitating the construction of valid Wald-style confidence intervals based on the asymptotic properties of the one-step bias-corrected estimator [69]. The precise specification of these estimators is as follows.

For the total effect, we have used the R package npcausal [70]. This package relies on the SuperLearner R package to specify models for fitting nuisance functions. We used:

(1) SL.earth, an implementation of multivariate adaptive regression splines [71];
(2) SL.glmnet, penalised regression with a generalised linear model and hyperparameter *α* = 1, i.e., *L*^1^-penalised or Least Absolute Shrinkage and Selection Operator (LASSO) regression, with default 10-fold cross-validation;
(3) SL.glm.interaction, generalised linear model with main terms and 2-way interactions;
(4) SL.xgboost, extreme gradient boosting (XGB) used with default parameters [72].

For the mediation effects NDE and NIE, we used the R package medoutcon [63]. This package instead relies on the sl3 R package [73], an implementation of the ensemble machine learning algorithm of [68], to specify models for fitting nuisance functions. We used:

(1) Lrnr_earth, an implementation of multivariate adaptive regression splines [71];
(2) Lrnr_glmnet, penalised linear regression with a generalised linear model and hyperparameters *α* = 1, i.e., *L*^1^-penalised or Least Absolute Shrinkage and Selection Operator (LASSO) regression, and default 3-fold cross-validation;
(3) Lrnr_glm_fast, a fast implementation of a generalised linear model used with main terms and 2-way interactions; and,
(4) Lrnr_lightgbm, a fast and memory-efficient implementation of extreme gradient boosting (XGB) models from the lightgbm R package [74], used with default parameters.

The estimation of NDE and NIE relies on the fitting of further nuisance functions for which we have used algorithms, such as the Highly Adaptive Lasso (HAL) [75, 76, 77], and parameter specifications recommended by medoutcon.

### 4.4. GO enrichment analysis

We performed Gene Ontology analysis [78, 79, 80] on the set of significant TE estimates (positive only, negative only, or all combined) obtained from the male, female or combined populations. For the background protein set, we used all 2,923 proteins measured in UKB.

We obtained significant results only for the set of proteins with a significant positive total effect in the female subset of the population at FDR < 0.05. The results are presented in Fig. S3. We used Rrvgo [80] to reduce redundancy of GO terms.

## Data Availability

No data has been produced. All data is from UK Biobank.

## Acknowledgements

This work was supported by a grant for PhD-level research to GLS from ME Research UK (SCIO charity number SCO36942). This research has been conducted using the UK Biobank Resource under Application Number 76173. Access to this data was funded by the National Institute for Health and Care Research (NIHR) and Medical Research Council (MRC) under grant number MC_PC_20005. AK was supported by a Langmuir Talent Development Fellowship from the Institute of Genetics and Cancer, and a philanthropic donation from Hugh and Josseline Langmuir. SB and AK acknowledge support of the UKRI AI programme, and the Engineering and Physical Sciences Research Council, for CHAI - EPSRC AI hub for Causality in Healthcare AI with real data [grant number EP/Y028856/1]. SB, AK and CPP are thankful to M. E. Khamseh for helpful discussions, and to Prof Jo Edwards, Charlie Hillier, Simon McGrath and members of the Science for ME forum for commenting on the initial preprint.

---

**Figure S1.**
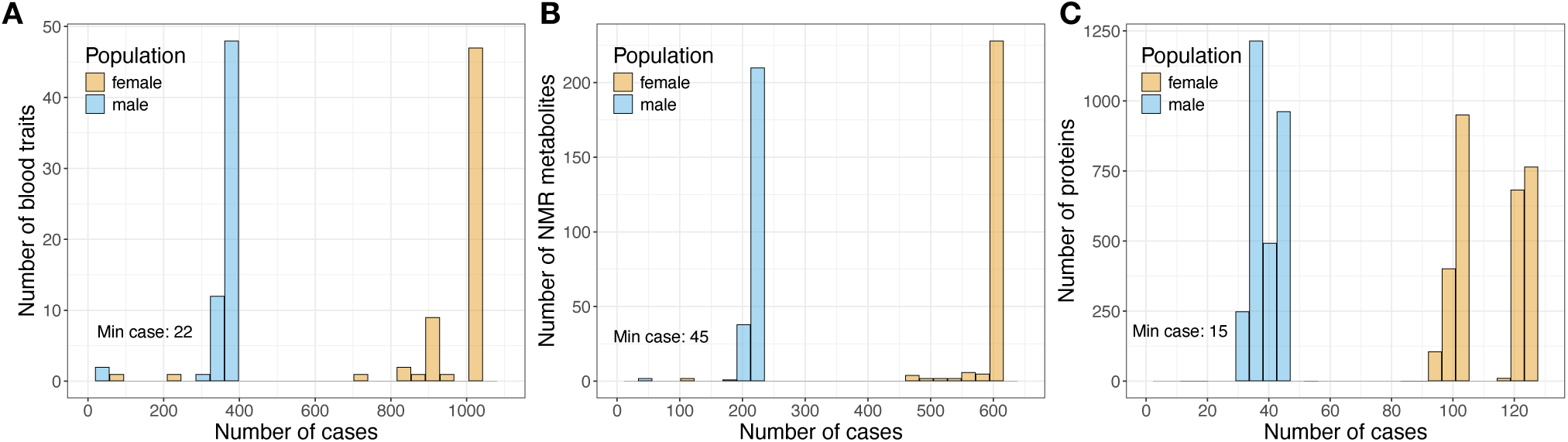
ME/CFS sample sizes for males and females, restricting to complete cases (individuals for whom a measurement is available). The minimum number of cases is indicated on each plot. (A) Blood traits, (B) NMR metabolites, (C) Proteomics. Neither of the two proteins with case sample size below 30 is significant after FDR correction. Full sample size data is provided as Supplementary Table 6.

**Figure S2.**
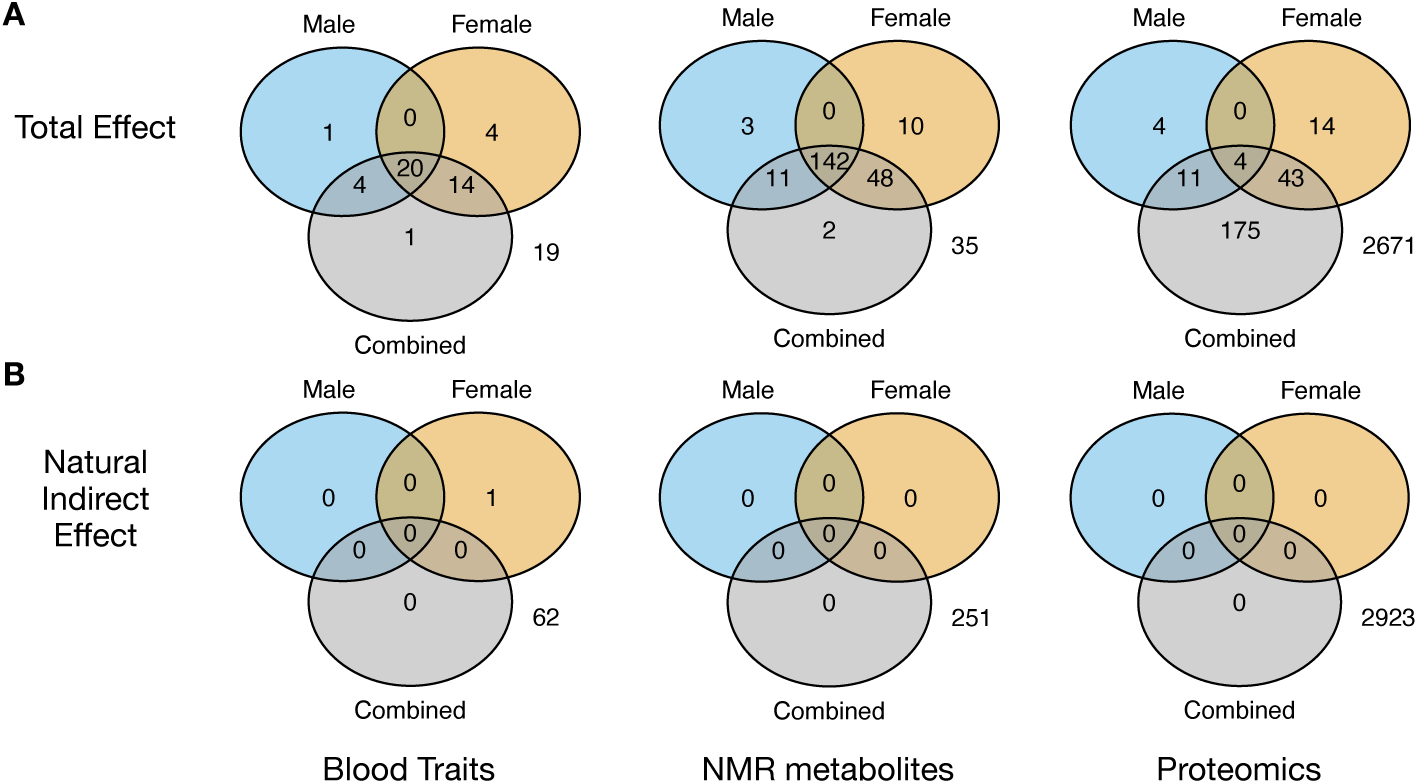
Venn diagrams displaying the number of significant findings in the males, females, combined and their intersection, mediator 874, for (A) total effect, and (B) NIE.

**Figure S3.**
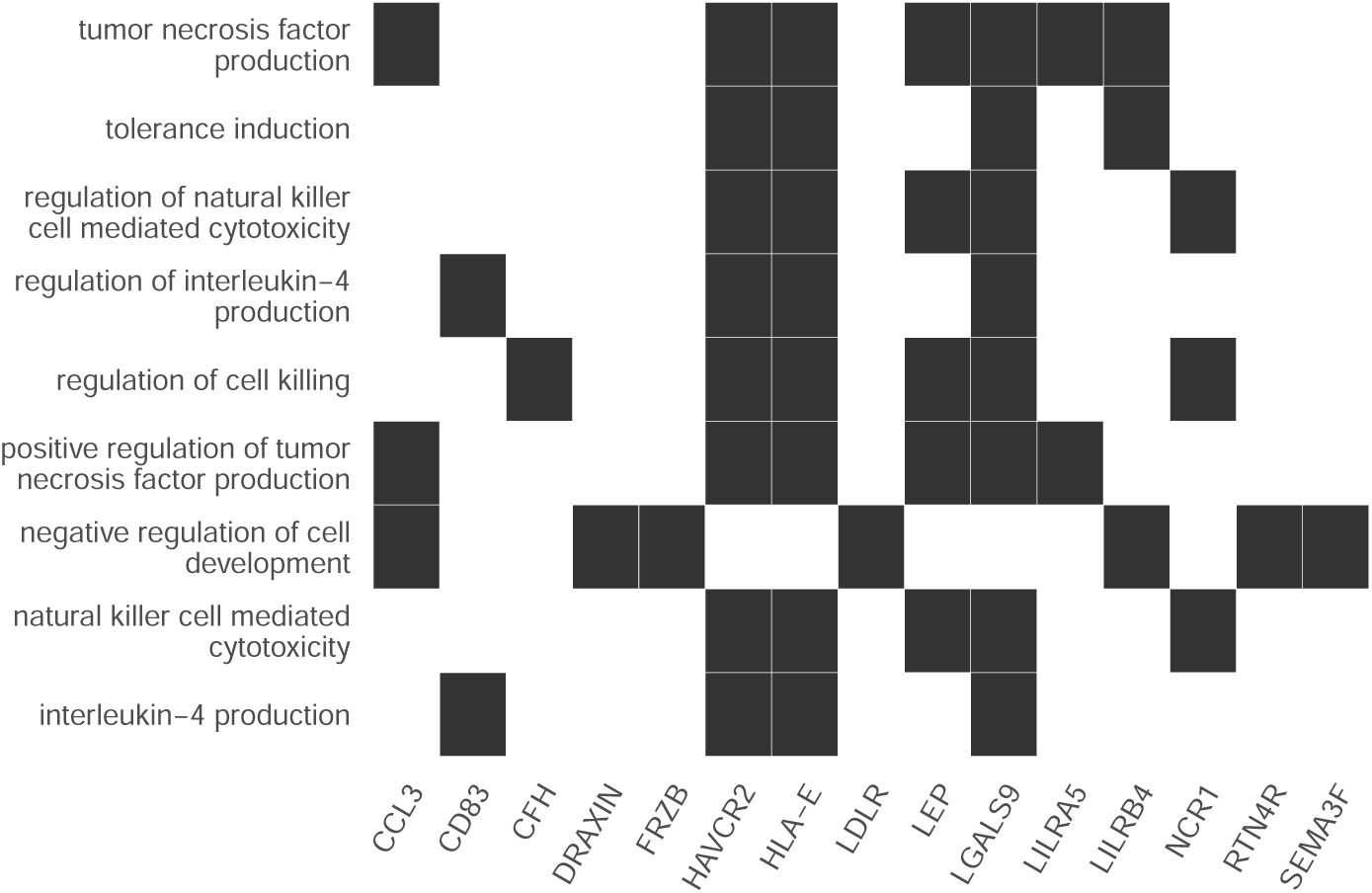
GO pathway enrichment [78] for proteins with a significant positive total effect for ME/CFS vs control, restricted to females only. This is the subset with maximal power for GO analysis. All effects are TE, *i.e.*, there are no significant NIE for proteins. We performed a similar pathway GO enrichment analysis for proteins with a significant positive total effect for ME/CFS vs control on the population of males and the combined dataset, as well as all significant negative total effects and all significant total effects on the female, male and combined populations. These resulted in no significant GO term enrichments at FDR< 0.05. All measured UKB proteins were used as background for the GO analyses.

**Figure S4.**
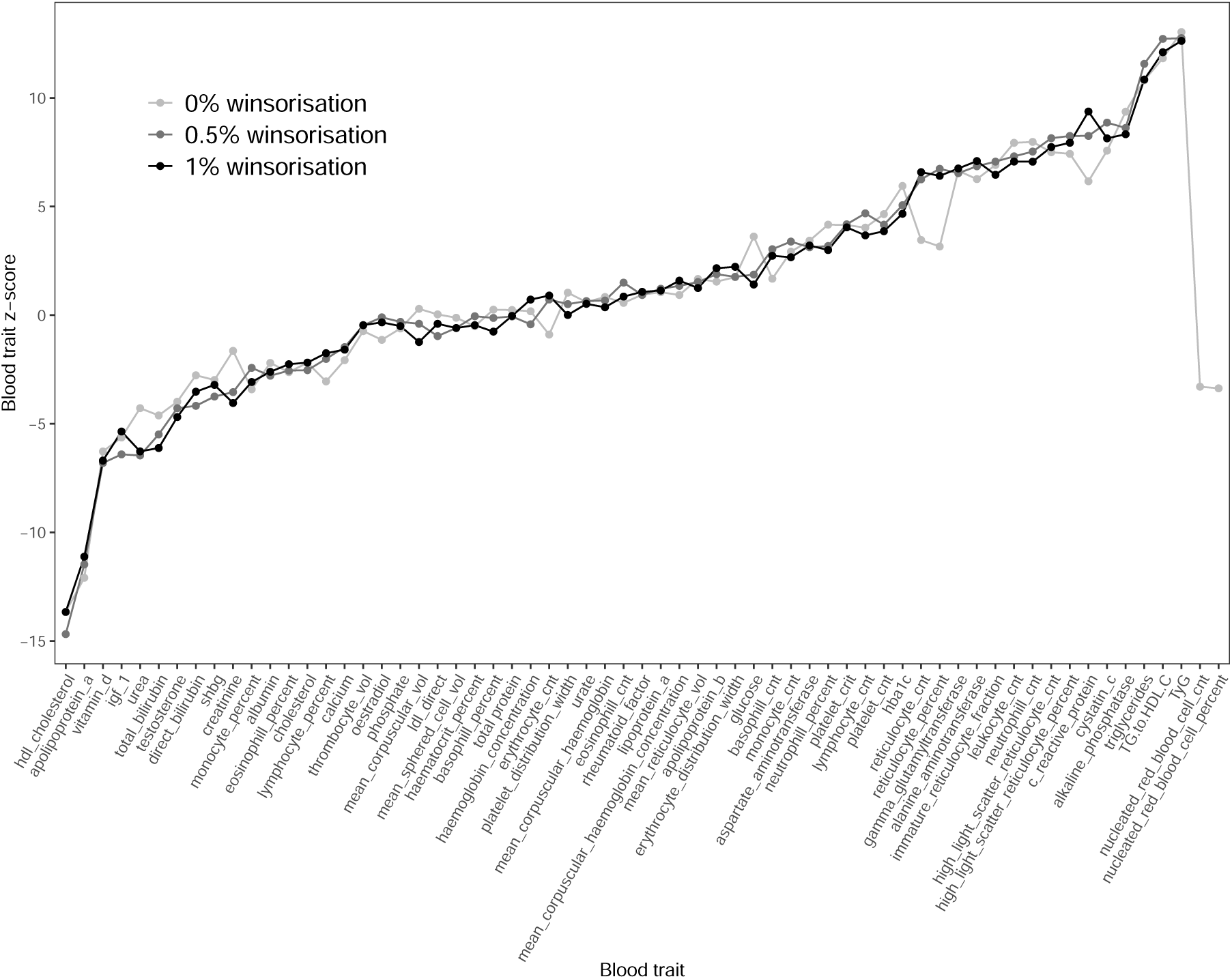
Significant blood traits are robust to winsorisation. The points represent total effect z-scores for blood traits in the combined female and male analysis. The three shades of grey represent different degrees of winsorisation of the original data, with cases and controls combined prior to winsorisation. Nucleated red blood cell count and percent are only estimable at 0% winsorisation because for 0.5% winsorisation the number of cases is *≤* 5. Fib4 and eGFR composite measures were not estimated for 0% winsorisation due to extreme values in control samples (*e.g.*, individuals with platelet counts close to 0).

## References

[1] Medicine I, Populations B, Syndrome C. Beyond myalgic encephalomyelitis/chronic fatigue syndrome: Redefining an illness; 2015.

[2] Cotler J, Holtzman C, Dudun C, Jason LA. A Brief Questionnaire to Assess Post-Exertional Malaise. Diagnostics. 2018;8(3).

[3] Myalgic encephalomyelitis (or encephalopathy)/chronic fatigue syndrome: diagnosis and management;. Accessed: 2023-08-15. https://www.nice.org.uk/guidance/ng206.

[4] Bretherick A, McGrath S, Devereux-Cooke A, Leary S, Northwood E, Redshaw A, et al. Typing myalgic encephalomyelitis by infection at onset: A DecodeME study. NIHR Open Research. 2023;3(20).

[5] Jason LA, Yoo S, Bhatia S. Patient perceptions of infectious illnesses preceding Myalgic Encephalomyelitis/Chronic Fatigue Syndrome. Chronic Illness. 2022;18(4):901–910. PMID: 34541918. Available from: 10.1177/17423953211043106.

[6] Tyson S, Stanley K, Gronlund TA, Leary S, Dean ME, Dransfield C, et al. Research priorities for myalgic encephalomyelitis/chronic fatigue syndrome (ME/CFS): the results of a James Lind alliance priority setting exercise. Fatigue: Biomedicine, Health & Behavior. 2022;10(4):200–211. Available from: 10.1080/21641846.2022.2124775.

[7] Maksoud R, Magawa C, Eaton-Fitch N, Thapaliya K, Marshall-Gradisnik S. Biomarkers for myalgic encephalomyelitis/chronic fatigue syndrome (ME/CFS): a systematic review. BMC Medicine. 2023;21(1):189.

[8] Huber K, Sunnquist M, Jason L. Latent class analysis of a heterogeneous international sample of patients with myalgic encephalomyelitis/chronic fatigue syndrome. Fatigue: Biomedicine, Health & Behavior. 2018 07;6:1–16.

[9] Silver A, Haeney M, Vijayadurai P, Wilks D, Pattrick M, Main CJ. The role of fear of physical movement and activity in chronic fatigue syndrome. Journal of Psychosomatic Research. 2002;52(6):485–493.

[10] Wessely S, David A, Butler S, Chalder T. Management of chronic (post-viral) fatigue syndrome. British Journal of General Practice. 1989;39(318):26–29.

[11] Moss-Morris R, Deary V, Castell B. Chapter 25 - Chronic fatigue syndrome. In: Barnes MP, Good DC, editors. Neurological Rehabilitation. vol. 110 of Handbook of Clinical Neurology. Elsevier; 2013. p. 303–314.

[12] Sharpe M. Cognitive behavior therapy for chronic fatigue syndrome. The American Journal of Medicine. 1995 2024/06/22;98(4):419–420.

[13] White P, Goldsmith K, Johnson A, Potts L, Walwyn R, DeCesare J, et al. Comparison of adaptive pacing therapy, cognitive behaviour therapy, graded exercise therapy, and specialist medical care for chronic fatigue syndrome (PACE): a randomised trial. The Lancet. 2011;377(9768):823–836. Available from: https://www.sciencedirect.com/science/article/pii/S0140673611600962.

[14] White PD. The role of physical inactivity in the chronic fatigue syndrome. Journal of Psychosomatic Research. 2000;49(5):283–284. Available from: https://www.sciencedirect.com/science/article/pii/S0022399900001951.

[15] for Health TNI, (NICE) CE. Myalgic encephalomyelitis (or encephalopathy)/chronic fatigue syndrome: diagnosis and management; 2021. Available from: https://www.nice.org.uk/guidance/ng206.

[16] Geraghty K, Jason L, Sunnquist M, Tuller D, Blease C, Adeniji C. The ‘cognitive behavioural model’ of chronic fatigue syndrome: Critique of a flawed model. Health Psychology Open. 2019;6(1):2055102919838907. PMID: 31041108. Available from: 10.1177/2055102919838907.

[17] of Medicine I. Beyond Myalgic Encephalomyelitis/Chronic Fatigue Syndrome: Redefining an Illness. Washington, DC: The National Academies Press; 2015. Available from: https://nap.nationalacademies.org/catalog/19012/beyond-myalgic-encephalomyelitischronic-fatigue-syndrome-redefining-an-illness.

[18] Bycroft C, Freeman C, Petkova D, Band G, Elliott LT, Sharp K, et al. The UK Biobank resource with deep phenotyping and genomic data. Nature. 2018;562(7726):203–209.

[19] Cairns R, Hotopf M. A systematic review describing the prognosis of chronic fatigue syndrome. Occupational Medicine. 2005 01;55(1):20–31. Available from: 10.1093/occmed/kqi013.

[20] Robins JM, Greenland S. Identifiability and Exchangeability for Direct and Indirect Effects. Epidemiology. 1992;3(2):143–155. Available from: http://www.jstor.org/stable/3702894.

[21] Pearl J. Direct and indirect effects. UAI’01. San Francisco, CA, USA: Morgan Kaufmann Publishers Inc.; 2001. p. 411–420.

[22] Benjamini Y, Hochberg Y. Controlling the False Discovery Rate: A Practical and Powerful Approach to Multiple Testing. Journal of the Royal Statistical Society Series B (Methodological). 1995;57(1):289–300. Available from: http://www.jstor.org/stable/2346101.

[23] Fritz J, Bjørge T, Nagel G, Manjer J, Engeland A, Häggström C, et al. The triglyceride-glucose index as a measure of insulin resistance and risk of obesity-related cancers. International Journal of Epidemiology. 2019 04;49(1):193–204. Available from: 10.1093/ije/dyz053.

[24] Won KB, Park EJ, Han D, Lee JH, Choi SY, Chun EJ, et al. Triglyceride glucose index is an independent predictor for the progression of coronary artery calcification in the absence of heavy coronary artery calcification at baseline. Cardiovascular Diabetology. 2020;19(1):34.

[25] Oliveri A, Rebernick RJ, Kuppa A, Pant A, Chen Y, Du X, et al. Comprehensive genetic study of the insulin resistance marker TG:HDL-C in the UK Biobank. Nature Genetics. 2024;56(2):212–221.

[26] Nagy-Szakal D, Barupal DK, Lee B, Che X, Williams BL, Kahn EJR, et al. Insights into myalgic encephalomyelitis/chronic fatigue syndrome phenotypes through comprehensive metabolomics. Scientific Reports. 2018;8(1):10056.

[27] Naviaux RK, Naviaux JC, Li K, Bright AT, Alaynick WA, Wang L, et al. Metabolic features of chronic fatigue syndrome. Proceedings of the National Academy of Sciences. 2016;113(37):E5472–E5480. Available from: https://www.pnas.org/doi/abs/10.1073/pnas.1607571113.

[28] Yamano E, Sugimoto M, Hirayama A, Kume S, Yamato M, Jin G, et al. Index markers of chronic fatigue syndrome with dysfunction of TCA and urea cycles. Scientific Reports. 2016;6(1):34990.

[29] Ghali A, Lacout C, Ghali M, Gury A, Beucher AB, Lozac’h P, et al. Elevated blood lactate in resting conditions correlate with post-exertional malaise severity in patients with Myalgic encephalomyelitis/Chronic fatigue syndrome. Scientific Reports. 2019;9(1):18817.

[30] Wang ZQ, Porreca F, Cuzzocrea S, Galen K, Lightfoot R, Masini E, et al. A Newly Identified Role for Superoxide in Inflammatory Pain. Journal of Pharmacology and Experimental Therapeutics. 2004;309(3):869–878. Available from: https://jpet.aspetjournals.org/content/309/3/869.

[31] Crawley SW, Shifrin DA, Grega-Larson NE, McConnell RE, Benesh AE, Mao S, et al. Intestinal Brush Border Assembly Driven by Protocadherin-Based Intermicrovillar Adhesion. Cell. 2014;157(2):433–446. Available from: https://www.sciencedirect.com/science/article/pii/S0092867414002153.

[32] Mödl B, Awad M, Zwolanek D, Scharf I, Schwertner K, Milovanovic D, et al. Defects in microvillus crosslinking sensitize to colitis and inflammatory bowel disease. EMBO reports. 2023;24(10):e57084. Available from: https://www.embopress.org/doi/abs/10.15252/embr.202357084.

[33] Triantafyllou G, Paschou S, Mantzoros C. Leptin and Hormones: Energy Homeostasis. Endocrinology & Metabolism Clinics of North America. 2016 09;45:633–45.

[34] Eaton-Fitch N, du Preez S, Cabanas H, Staines D, Marshall-Gradisnik S. A systematic review of natural killer cells profile and cytotoxic function in myalgic encephalomyelitis/chronic fatigue syndrome. Systematic Reviews. 2019;8(1):279.

[35] Taylor J, Tibshirani RJ. Statistical learning and selective inference. Proceedings of the National Academy of Sciences. 2015;112(25):7629–7634.

[36] Faro M, Sàez-Francás N, Castro-Marrero J, Aliste L, Fernández de Sevilla T, Alegre J. Gender Differences in Chronic Fatigue Syndrome. Reumatología Clínica (English Edition). 2016;12(2):72–77.

[37] Jopson L, Dyson JK, Jones DEJ. Understanding and Treating Fatigue in Primary Biliary Cirrhosis and Primary Sclerosing Cholangitis. Clinics in Liver Disease. 2016;20(1):131–142. Advances in Cholestatic Liver Diseases. Available from: https://www.sciencedirect.com/science/article/pii/S1089326115000793.

[38] Cohen J. Statistical Power Analysis for the Behavioral Sciences. Taylor & Francis; 2013. Available from: https://books.google.co.uk/books?id=cIJH0lR33bgC.

[39] Fry A, Littlejohns TJ, Sudlow C, Doherty N, Adamska L, Sprosen T, et al. Comparison of Sociodemographic and Health-Related Characteristics of UK Biobank Participants With Those of the General Population. American Journal of Epidemiology. 2017 06;186(9):1026–1034. Available from: 10.1093/aje/kwx246.

[40] Samms GL, Ponting CP. Unequal access to diagnosis of myalgic encephalomyelitis in England. medRxiv. 2024;Available from: https://www.medrxiv.org/content/early/2024/02/01/2024.01.31.24302070.

[41] Shankar V, Wilhelmy J, Curtis EJ, Michael B, Cervantes L, Mallajosyula VA, et al. Oxidative Stress is a shared characteristic of ME/CFS and Long COVID. bioRxiv. 2024;.

[42] Xiong R, Fleming E, Caldwell R, Vernon SD, Kozhaya L, Gunter C, et al. BioMapAI: Artificial Intelligence Multi-Omics Modeling of Myalgic Encephalomyelitis / Chronic Fatigue Syndrome. bioRxiv. 2024;.

[43] Stamatakis E, Owen KB, Shepherd L, Drayton B, Hamer M, Bauman AE. Is Cohort Representativeness Passé? Poststratified Associations of Lifestyle Risk Factors with Mortality in the UK Biobank. Epidemiology. 2021;32(2).

[44] Davis KAS, Coleman JRI, Adams M, Allen N, Breen G, Cullen B, et al. Mental health in UK Biobank –development, implementation and results from an online questionnaire completed by 157 366 participants: a reanalysis. BJPsych Open. 2020;6(2):e18.

[45] Elliott P, Peakman oboUB Tim C. The UK Biobank sample handling and storage protocol for the collection, processing and archiving of human blood and urine. International Journal of Epidemiology. 2008 04;37(2):234–244. Available from: 10.1093/ije/dym276.

[46] UK Biobank biochemistry assay quality procedures;. Accessed: 2024-07-28. https://biobank.ndph.ox.ac.uk/showcase/refer.cgi?id=5636.

[47] UK Biobank companion document for serum biomarker data;. Accessed: 2024-07-28. https://biobank.ndph.ox.ac.uk/showcase/refer.cgi?id=1227.

[48] Che B, Zhong C, Zhang R, Pu L, Zhao T, Zhang Y, et al. Triglyceride-glucose index and triglyceride to high-density lipoprotein cholesterol ratio as potential cardiovascular disease risk factors: an analysis of UK biobank data. Cardiovascular Diabetology. 2023;22(1):34.

[49] Si S, Li J, Li Y, Li W, Chen X, Yuan T, et al. Causal Effect of the Triglyceride-Glucose Index and the Joint Exposure of Higher Glucose and Triglyceride With Extensive Cardio-Cerebrovascular Metabolic Outcomes in the UK Biobank: A Mendelian Randomization Study. Frontiers in Cardiovascular Medicine. 2021;7. Available from: https://www.frontiersin.org/articles/10.3389/fcvm.2020.583473.

[50] Cordero A, Alegria-Ezquerra E. TG/HDL ratio as surrogate marker for insulin resistance; 2009. Available from: https://www.escardio.org/Journals/E-Journal-of-Cardiology-Practice/Volume-8/TG-HDL-ratio-as-surrogate-marker-for-insulin-resistance.

[51] Simental-Mendía LE, Rodríguez-Morán M, Guerrero-Romero F. The product of fasting glucose and triglycerides as surrogate for identifying insulin resistance in apparently healthy subjects. Metabolic syndrome and related disorders. 2008 December;6(4):299—304. Available from: 10.1089/met.2008.0034.

[52] VanderWeele TJ. Mediation Analysis: A Practitioner’s Guide [Journal Article]. Annual Review of Public Health. 2016;37(Volume 37, 2016):17–32. Available from: https://www.annualreviews.org/content/journals/10.1146/annurev-publhealth-032315-021402.

[53] Nguyen TQ, Schmid I, Stuart EA. Clarifying causal mediation analysis for the applied researcher: Defining effects based on what we want to learn. Psychological Methods. 2021 Apr;26(2):255–271. Available from: 10.1037/met0000299.

[54] Hejazi NS, van der Laan MJ, Janes HE, Gilbert PB, Benkeser DC. Efficient Nonparametric Inference on the Effects of Stochastic Interventions under Two-Phase Sampling, with Applications to Vaccine Efficacy Trials. Biometrics. 2020 09;77(4):1241–1253. Available from: 10.1111/biom.13375.

[55] Benkeser D, Montefiori DC, McDermott AB, Fong Y, Janes HE, Deng W, et al. Comparing antibody assays as correlates of protection against COVID-19 in the COVE mRNA-1273 vaccine efficacy trial. Science Translational Medicine. 2023;15(692):eade9078. Available from: https://www.science.org/doi/abs/10.1126/scitranslmed.ade9078.

[56] Huang Y, Hejazi NS, Blette B, Carpp LN, Benkeser D, Montefiori DC, et al. Stochastic interventional vaccine efficacy and principal surrogate analyses of antibody markers as correlates of protection against symptomatic COVID-19 in the COVE mRNA-1273 trial. Viruses. 2023;15(10):2029.

[57] Hejazi NS, Shen X, Carpp LN, Benkeser D, Follmann D, Janes HE, et al. Stochastic interventional approach to assessing immune correlates of protection: Application to the COVE mRNA-1273 vaccine trial. International Journal of Infectious Diseases. 2023;137:28–39.

[58] Rudolph KE, Díaz I, Hejazi NS, van der Laan MJ, Luo SX, Shulman M, et al. Explaining differential effects of medication for opioid use disorder using a novel approach incorporating mediating variables. Addiction. 2021;116(8):2094–2103. Available from: https://onlinelibrary.wiley.com/doi/abs/10.1111/add.15377.

[60] Hejazi NS, Rudolph KE, Van Der Laan MJ, Díaz I. Nonparametric causal mediation analysis for stochastic interventional (in)direct effects. Biostatistics. 2022 02;24(3):686–707. Available from: 10.1093/biostatistics/kxac002.

[60] Rudolph KE, Gimbrone C, Díaz I. Helped into Harm: Mediation of a Housing Voucher Intervention on Mental Health and Substance Use in Boys. Epidemiology. 2021;32(3).

[61] Menkir TF, Citarella B, Sigfrid L, Doshi Y, Reyes F, Calvache JA, et al. Modeling the relative influence of socio-demographic variables on post-acute COVID-19 quality of life: an application to settings in Europe, Asia, Africa, and South America. 2024;.

[62] Díaz I, Hejazi NS, Rudolph KE, van der Laan MJ. Non-parametric efficient causal mediation with intermediate confounders. Biometrika. 2020;108(3):627–641. Available from: https://arxiv.org/abs/1912.09936.

[63] Hejazi NS, Rudolph KE, Díaz I. medoutcon: Nonparametric efficient causal mediation analysis with machine learning in R. Journal of Open Source Software. 2022;Available from: 10.21105/joss.03979.

[64] Hejazi NS, Díaz I, Rudolph KE. medoutcon: Efficient natural and interventional causal mediation analysis; 2022. R package version 0.1.6. Available from: https://github.com/nhejazi/medoutcon.

[65] Rubin DB. Bayesian Inference for Causal Effects: The Role of Randomization. The Annals of Statistics. 1978;6(1):34–58. Available from: http://www.jstor.org/stable/2958688.

[66] Rubin DB. Randomization Analysis of Experimental Data: The Fisher Randomization Test Comment. Journal of the American Statistical Association. 1980;75(371):591–593. Available from: http://www.jstor.org/stable/2287653.

[67] Robins JM, Richardson TS. Alternative graphical causal models and the identification of direct effects. In: Shrout P, Keyes K, Ornstein K, editors. Causality and psychopathology: Finding the determinants of disorders and their cures. Oxford: Oxford University Press; 2011.

[68] van der Laan MJ, Polley EC, Hubbard AE. Super learner. Stat Appl Genet Mol Biol. 2007;6:Art. 25, 23.

[69] Bickel PJ, Klaassen CAJ, Ritov Y, Wellner JA. Efficient and Adaptive Estimation for Semiparametric Models. Johns Hopkins series in the mathematical sciences. Springer New York; 1998. Available from: https://books.google.co.uk/books?id=lSnTm6SC_SMC.

[70] Kennedy E. npcausal: Nonparametric causal inference methods; 2021. R package version 0.1.1. Available from: https://github.com/ehkennedy/npcausal.

[71] Friedman JH. Multivariate Adaptive Regression Splines. The Annals of Statistics. 1991;19(1):1–67.

[72] Chen T, Guestrin C. XGBoost: A Scalable Tree Boosting System. In: Proceedings of the 22nd ACM SIGKDD International Conference on Knowledge Discovery and Data Mining. KDD ’16. New York, NY, USA: Association for Computing Machinery; 2016. p. 785–794. Available from: 10.1145/2939672.2939785.

[72] Coyle JR, Hejazi NS, Malenica I, Phillips RV, Sofrygin O. sl3: Super Machine Learning with pipelines;. R package. Available from: https://github.com/tlverse/sl3.

[74] Ke G, Meng Q, Finley T, Wang T, Chen W, Ma W, et al. LightGBM: A Highly Efficient Gradient Boosting Decision Tree. In: Guyon I, Luxburg UV, Bengio S, Wallach H, Fergus R, Vishwanathan S, et al., editors. Advances in Neural Information Processing Systems. vol. 30. Curran Associates, Inc.; 2017. Available from: https://proceedings.neurips.cc/paper_files/paper/2017/file/6449f44a102fde848669bdd9eb6b76fa-Paper.pdf.

[75] van der Laan MJ. A generally efficient targeted minimum loss based estimator based on the highly adaptive lasso. The international journal of biostatistics. 2017;13(2).

[76] Hejazi NS, Coyle JR, van der Laan MJ. hal9001: Scalable highly adaptive lasso regression in R. Journal of Open Source Software. 2020 9;5(53):2526. Available from: 10.21105/joss.02526.

[77] Coyle JR, Hejazi NS, Phillips RV, van der Laan LWP, van der Laan MJ. hal9001: The scalable highly adaptive lasso;. R package. Available from: https://CRAN.R-project.org/package=hal9001.

[78] Ashburner M, Ball CA, Blake JA, Botstein D, Butler H, Cherry JM, et al. Gene Ontology: tool for the unification of biology. Nature Genetics. 2000;25(1):25–29.

[79] Aleksander SA, Balhoff J, Carbon S, Cherry JM, Drabkin HJ, Ebert D, et al. The Gene Ontology knowledgebase in 2023. Genetics. 2023 03;224(1):iyad031. Available from: 10.1093/genetics/iyad031.

[80] Sayols S. rrvgo: a Bioconductor package for interpreting lists of Gene Ontology terms. microPublication Biology. 2023;Available from: https://www.micropublication.org/journals/biology/micropub-biology-000811.

