## Supplementary Table captions for "Replicated blood-based biomarkers for Myalgic Encephalomyelitis not explicable by inactivity"

List of Supplementary Tables

**Supplementary Table 1:** ME/CFS Primary Care record (CTV3 or Read v2 code)

**Supplementary Table 2:** Robust blood trait biomarkers

**Supplementary Table 3:** Blood trait biomarkers for TE, NDE, NIE

with mediator 874 and super learner

**Supplementary Table 4:** NMR biomarkers for TE, NDE, NIE with mediator 874

**Supplementary Table 5:** Proteomics biomarkers for TE, NDE, NIE with mediator 874

**Supplementary Table 6:** Number of case and controls samples used in each TE estimate, for BT, NMR and proteomics

**Supplementary Table 7:** Blood trait biomarkers sensitivity analysis for NDE and NIE with mediators 884 and 894, and super learner

**Supplementary Table 8:** Blood trait biomarkers sensitivity analysis for TE with winsorisation (0%, 0.5%, 1% extreme values removed), and super learner

**Supplementary Table 9:** Blood trait biomarkers sensitivity analysis for TE, NDE, NIE with mediator 874 and no super learner

**Supplementary Table 10:** Blood trait biomarkers case and control definition sharpening analysis for TE, NDE, NIE with mediator 874 and no super learner

**Supplementary Table 11:** Cohen’s D estimates for BT, NMR, and proteomics
